# Strategies to support safe wandering in care homes for older adults – what works, for whom, and in which circumstances?: A realist synthesis

**DOI:** 10.1101/2025.07.18.25331765

**Authors:** Emma S. Hock, Bryony Waters-Harvey, Alys Wyn Griffiths, Emily Fisher, Tamara Backhouse, Iria Cunha, Sion Scott, Clarissa Giebel, Liz Jones, Jignasa Mehta, Karen Spilsbury, Andrew Booth, Reena Devi, Mary Gemma Cherry

**Author notes:** Corresponding author: Mary Gemma Cherry. denotes joint last author (Reena Devi, Mary Gemma Cherry).

## Abstract

**Objectives:** Wandering is a common behaviour among people with dementia living in care homes, driven by various factors such as enjoyment, a sense of purpose, lifelong habits, and social interaction. These elements can bring both physical and mental benefits, highlighting the need for strategies that enable safe wandering while respecting individual autonomy. This realist synthesis aimed to explore these strategies and the conditions under which they lead to successful outcomes.

**Methods:** This realist synthesis involved scoping the literature to develop initial theoretical explanations for how different strategies could support safe wandering. From this literature, we developed context-mechanism-outcome configurations, which we combined into initial programme theories (IPTs). Systematic searches were then used to test and refine these programme theories. Studies were prioritised for inclusion based on criteria of relevance and richness. We extracted data pertinent to the IPTs and documented relevance, richness, and rigour. We synthesised data into five refined programme theories. At each stage of the process, we collaborated with stakeholders to develop and validate the strategies.

**Results:** The review included 79 evidence sources, leading to five refined programme theories. 1) **Personalised Care**: Emphasising the importance of staff practicing person-centred care by understanding residents, their reasons for wandering, and their life histories. 2) **Monitoring**: Effective monitoring requires good visual access or technological solutions that enable staff to observe residents, and detect when residents need support to walk while also enabling resident freedom and independence. 3) **Navigation**: Navigation is facilitated by dementia-friendly design features and environmental cues, which help minimise the challenges residents face due to diminished orientation and wayfinding abilities. 4) **Managing access**: Involving balancing residents’ safety and autonomy. Strategies may include restricting access to unsafe areas by locking doors or using technology and camouflage, while ensuring access to safe spaces. 5) **Hydration and nutrition**: Hydration and nutrition (e.g., suitable snacks) is provided to prevent weight loss for residents who wander and may not stay seated during meals. These theories provide insight into supporting safe wandering, leading to improved wellbeing for both residents and staff, enhanced safety and autonomy for residents, and reduced staff anxiety.

**Discussion:** Strategies that create a supportive environment, provide physical assistance, and support hydration and nutrition enabled residents to wander safely. Identified strategies improved wellbeing for both residents and staff. However, the same strategies also led to ethical concerns around digital monitoring, deception, and access restrictions.

**Study registration:** The protocol was registered with PROSPERO (CRD2024559085).

**Highlights:** *What is known:*

- There are many barriers to supporting people to wander safely in care homes despite the physical and mental health benefits this may bring
- There is little knowledge of which strategies work for whom and in what circumstances

*What this paper adds:*

- We identified and synthesised strategies to promote safe wandering in care homes, relating to personalised care, monitoring, navigation, managing access, and hydration and nutrition
- Residents’ independence and autonomy, along with the well-being of both residents and staff, are enhanced when residents can wander safely
- Some strategies raise ethical concerns related to digital monitoring, deception, and access restrictions

## Introduction

Around 60% of people living with dementia will wander whilst residing in care homes (1), and wandering often precedes individuals transitioning into care homes (2). Wandering is characterised by walking slowly in a relaxed way, often without a specific purpose or direction (3) and can be challenging for carers and family members to manage due to risks associated with wandering (4–6), such as falls, dehydration, malnutrition, and mortality (7). There is no consensus on the definition of wandering in dementia care (8), although significant debate surrounds the term (see footnote^1^), but characteristics associated with the term include a high frequency and repetition of walking (9). Care home staff may interpret frequent walking or being lost, restless, unable to sit down, hyperactive, and/or confused as being associated with wandering (4, 10). However, whilst the purpose and aim may not be understood by others (11). People with dementia give diverse reasons for wandering, including enjoyment, purpose, lifelong habits and socialisation (12).

Care home staff may find it stressful to support people who demonstrate wandering behaviours (13), and as a result, many families and staff prioritise safety over the right to freedom and independence. Historically, restrictive measures, including physical and chemical restraints, have been used to prevent wandering (14, 15). However, restrictive measures raise ethical concerns and can lead to adverse effects such as anxiety, falls, pressure sores and increased morbidity and mortality (16–19). Restraints are still commonly used, such as bed rails and preventing residents from rising from their chairs; although these are not typically perceived by staff as restraints (20, 21). Psychosocial interventions such as exercise, music therapy, and aromatherapy (14, 16) are commonly implemented. However, these strategies also aim, to some extent, to prevent wandering rather than promote autonomy and movement.

Wandering is sometimes not fully understood by staff members and families, leading to them dismissing its benefits due to perceived risks (14). However, the opportunity to walk around care homes is beneficial for older adults both physically and mentally, provided that they are supported to wander safely (12). Wandering provides physical activity, which in turn can improve circulation, reduce deconditioning, foster a sense of independence and autonomy (12, 14, 22), and reduce anxiety or conflicts between residents (14). Staff need to manage a balance between promoting safe wandering and managing its associated risks (8).

To value and promote wandering, effective strategies that enable safe wandering while maintaining individual autonomy should need to be considered. To determine what works, for whom, in what contexts and why, a realist review was conducted to explore:

1) How do strategies to support safe wandering for older adults in care homes work, and why?
2) In what circumstances can successful outcomes be expected for these strategies?

## Methods

A realist synthesis approach (23) was used. The protocol was registered with PROSPERO (registration number CRD2024559085). Reporting followed the Realist And MEta-narrative Evidence Syntheses: Evolving Standards (RAMESES) publication standards (24).

### Initial programme theory development

We undertook scoping literature searches in August 2023 across PubMed, PsycInfo and CINAHL databases, combining terms for ‘wandering’, ‘care homes’ and ‘dementia’, supplemented with hand searches of reference lists. Relevant studies were qualitative and explored any aspect of wandering in care homes for people with dementia. We identified 17 evidence sources, including empirical papers and reviews, which were dual-extracted to identify initial context-mechanism-outcome configurations (CMOCs). We also consulted lay advisory group members (friends and family of people who live/have lived in care homes), whose experiences contributed to CMOC generation.

We categorised the 287 initial CMOCs before refining these through discussion into 11 initial programme theories (IPTs), focusing on common mechanisms related to promoting safe wandering. Stakeholders (care home professionals, architects, and family/friends) completed an online or paper survey to rank their top five priority IPTs. We listed the IPTs in order of rated importance and prioritised the top five endorsed IPTs for further testing and refining, following consultation with stakeholders who highlighted the importance of those theories.

### Searches

An initial broad search for evidence to test the IPTs was conducted in PubMed, Cochrane Library, PsycInfo, CINAHL, ProQuest Dissertations & Theses Global, Scopus, and the NIHR Journals Library, using Boolean operators to combine terms relating to ‘wandering’ ‘care homes’ and ‘dementia’. Initial searches were limited to items published between 2005 and 2024 (for pragmatic reasons) and published in English (Supplement 1).

This broad search was followed by targeted searches focused on the five prioritised IPTs (Supplement 1). Full text searches were undertaken using Litsense and Scite, with combinations of terms and specific phrases from the IPTs plus ‘wandering’ and ‘dementia’. The most rich and relevant article that included evidence relating to most IPTs (25) was entered into Scite and Google Scholar, and citations and ‘related articles’ were examined. Relevant websites (identified through stakeholder consultation) were searched systematically via menu headings and by searching key terms relating to wandering and, where relevant, dementia (Supplement 1). One reviewer [redacted] conducted and documented grey literature searches (26).

### Study selection

Two reviewers [redacted] used Rayyan to double-screen titles and abstracts from the initial broad search. Disagreements and queries were resolved by discussion with [redacted]. [Redacted] double-screened full texts of papers included at the abstract stage, focusing on relevance and richness (Box 1). The same approach was used to screen, review, and select grey literature full texts.

##### Box 1: Definition of relevance, richness and rigour (27)

**Relevance:** The degree to which the source could contribute to theory building/testing, particularly in terms of descriptive data for one or more of our IPTs, and coding for dyads/triads (links between aspects of our IPTs)

**Richness:** The degree of ‘conceptual richness’ and/or ‘contextual thickness’ (sufficient detail on theoretical and conceptual development and contextual detail

**Rigour:** The methodological quality of the source, including representativeness and quality of contribution to the CMOC.

One reviewer [redacted] who conducted the IPT-focused searches, screened relevant extracts from each paper iteratively within the search results while searching, as Scite and Litsense present records in order of relevance. Potentially relevant sources were checked against the Rayyan database and, if new, full texts were screened.

Following the study selection process, it was clear that many sources reported data relevant to two additional IPTs from our initial list. Therefore, we synthesised these, bringing the total number of IPTs synthesised to seven.

### Data extraction

Two reviewers [redacted] extracted data (study sample, methods, setting and care home context). Reviewers categorised IPT-related data by each relevant IPT at extraction, and logged judgements about relevance, richness and rigour (Box 1).

#### Synthesis

The initial unit for synthesis was the IPT. [redacted] synthesised data for each of the seven IPTs by reporting on contexts and mechanisms in relation to the outcomes of strategies used to support safe wandering (checked by [redacted]). Two reviewers [redacted] summarised the synthesis for review by the lay advisory group.

During the synthesis, we identified several common mechanisms and contexts, and developed an overarching programme theory to demonstrate links between them. Within this overarching theory, we identified natural groupings within which distinct concepts could be arranged. These resembled the IPTs but were not exactly matching. We then grouped findings within these distinct concepts and synthesised them into five refined programme theories.

#### Sense-checking conversations

After synthesising the data, we carried out ‘sense-checking’ of our findings with care home staff through one-on-one conversations and focus groups. We presented the review methodology and lay summaries of each refined programme theory to staff, who provided their reflections and feedback on whether our findings aligned with their experiences.

## Results

### Programme theory development

The original list of IPTs is provided in Figure 1, along with the prioritised IPTs, which were:

- Staff detect when residents need support (IPT3)
- Staff understanding of why each resident wanders (IPT4)
- Residents can’t access dangerous spaces (IPT7)
- Residents are supported physically (IPT9)
- Residents are adequately nourished and/or hydrated (IPT10)

And two additional IPTs with a large volume of evidence:

- Residents can orient themselves and find their way (IPT6)
- Residents can access spaces (IPT8)

**Figure 1:**
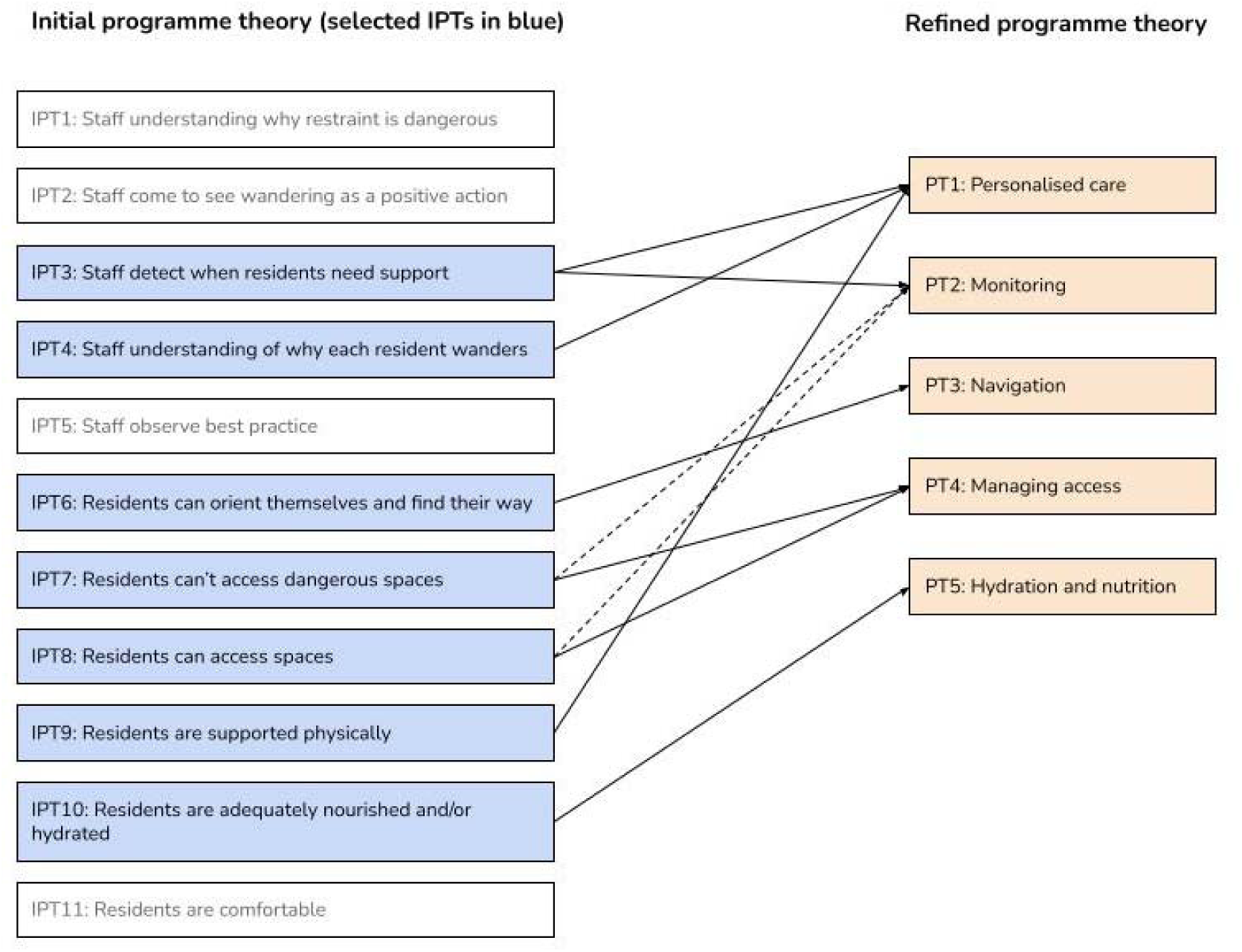
Programme theory refinement (selected IPTs are in blue, solid arrows represent a major contribution to refined PTs, dashed arrow represents a minor contribution to refined PTs)

### Volume of evidence

Of 2875 records retrieved through database searches, 155 full texts were screened and 57 were included. Of 5205 records retrieved through iterative IPT searches, 45 full texts were screened and 19 were included. Of 251 potentially relevant grey literature sources examined, three were included. In total, 79 evidence sources were included in the realist synthesis (Figure 2).

**Figure 2:**
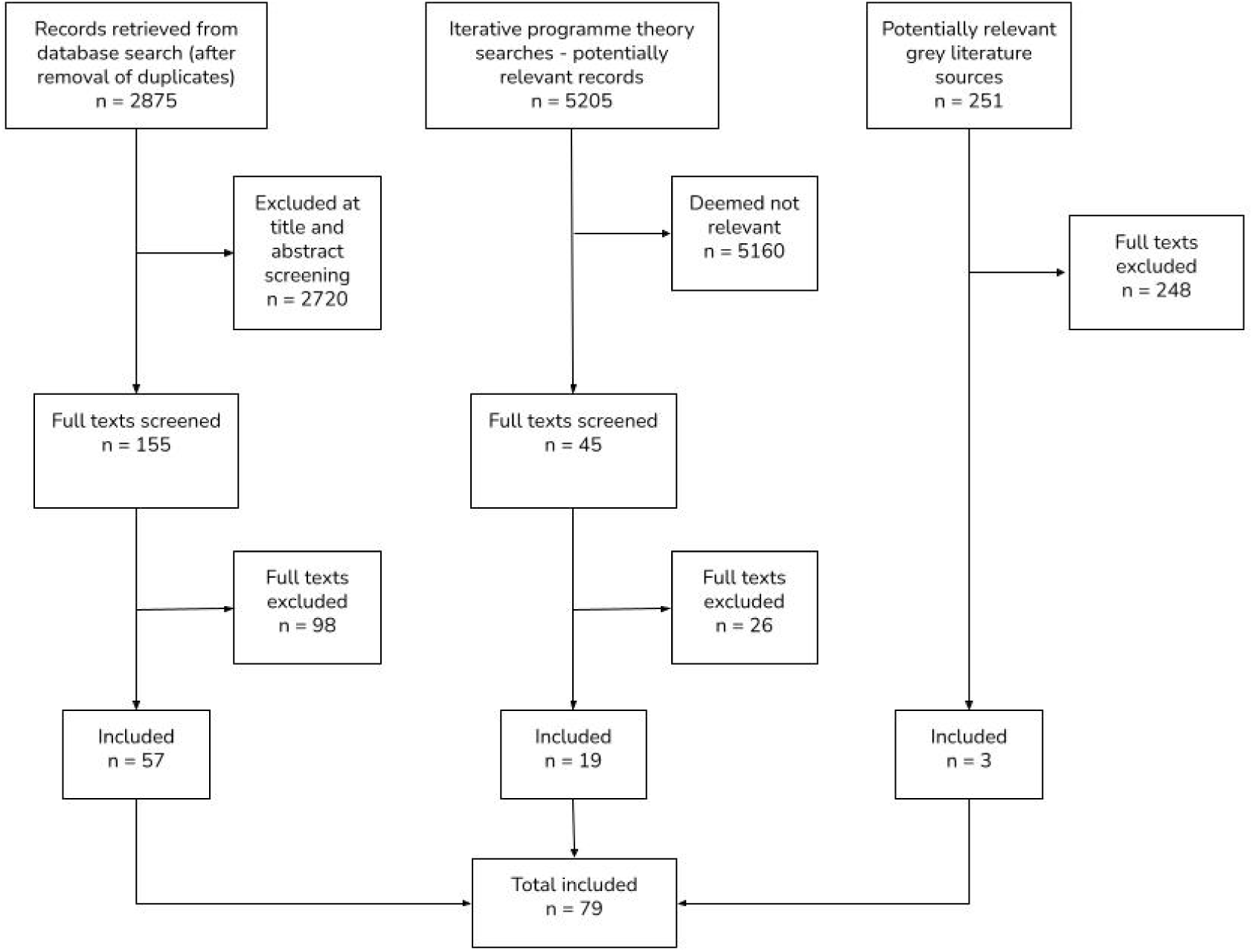
PRISMA diagram of study selection

Characteristics of the included evidence sources are presented in Table 1 and relevance, richness and rigour are further detailed in Supplement 2. Evidence sources were published between 1987 and 2025, from a range of countries. There were 18 qualitative studies, 17 quantitative, eight mixed methods, four case studies, seven systematic reviews, one meta-review, 11 reviews of other types, nine commentaries, and four grey literature sources. Evidence was generated in diverse settings including general care settings such as care homes, care communities, long-term care facilities and assisted living facilities, and those providing specialised support, such as nursing homes, dementia care units, special care units, psychiatric units, and related settings (e.g., a day centre, hospital, the community). Of the included evidence sources, 45 (57%) were considered trustworthy (rated high or moderate for rigour), and 34 (43%) were classed as untrustworthy (rated low for rigour). Twenty-seven (34%) were rated high, 43 (54%) were rated moderate, and 9 (11%) were rated as being low relevance. Ten (13%) were rated high, 49 (62%) were rated moderate and 20 (25%) were rated low richness (Table 1).

**Table 1:**
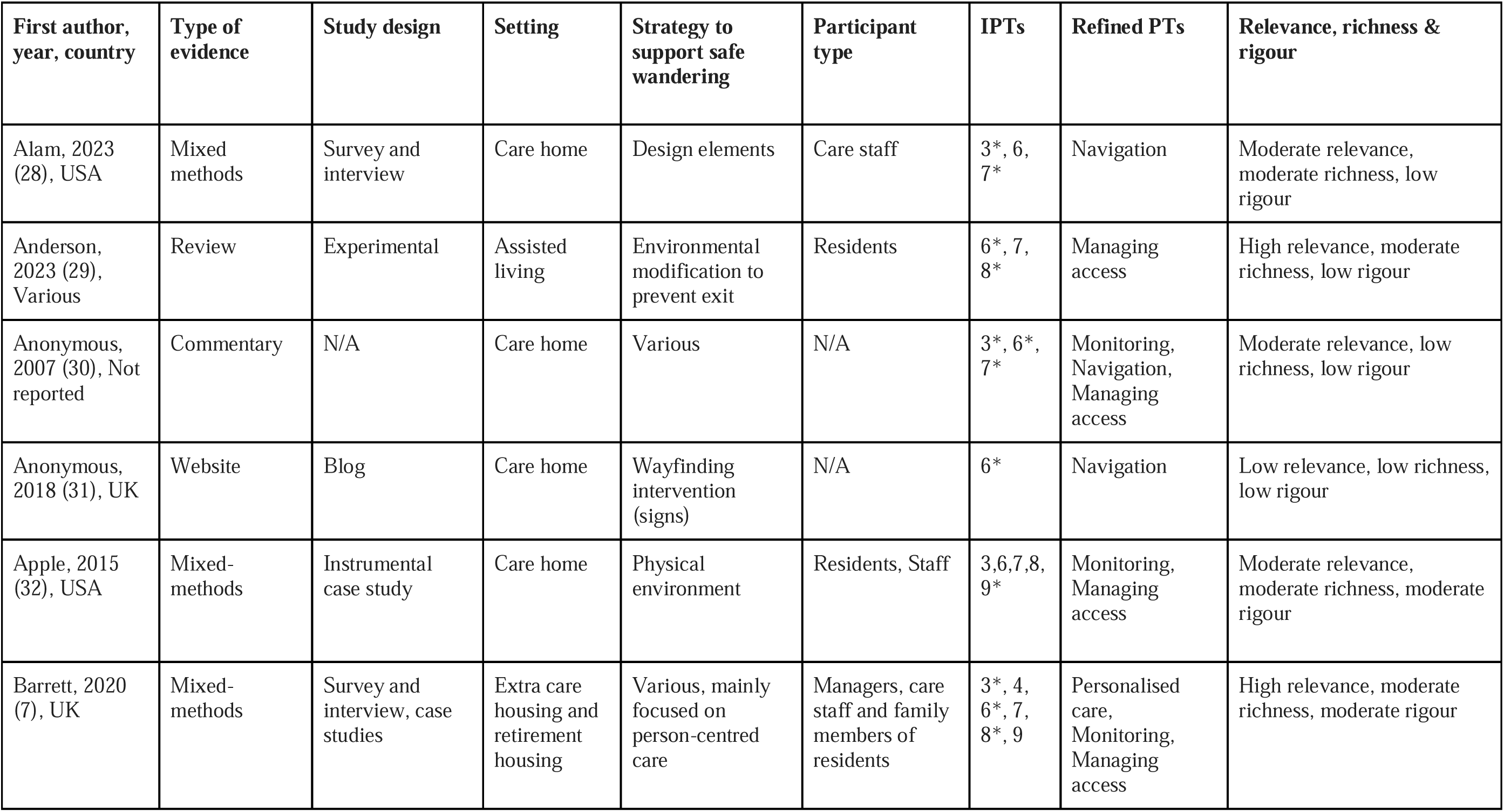
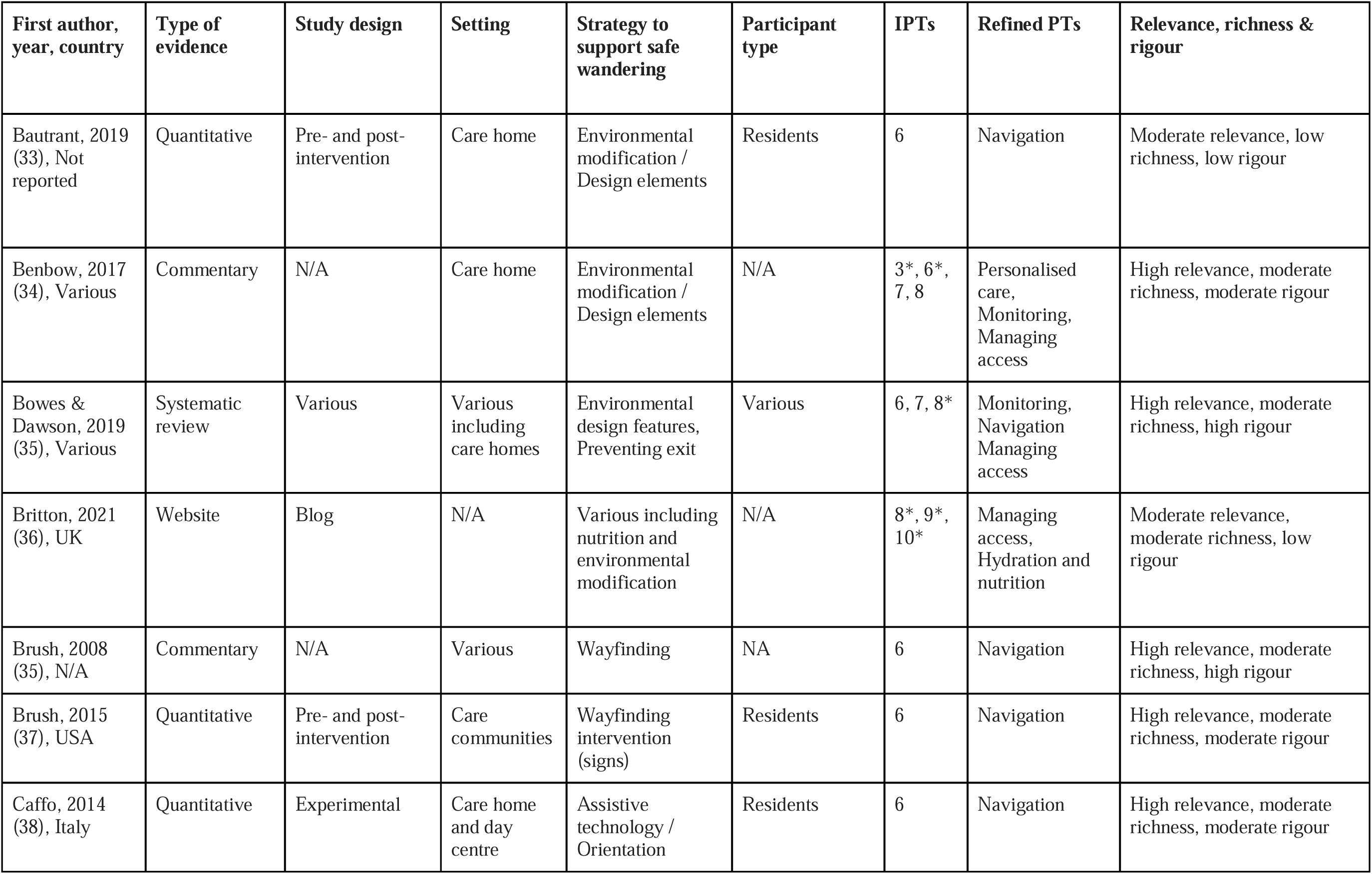
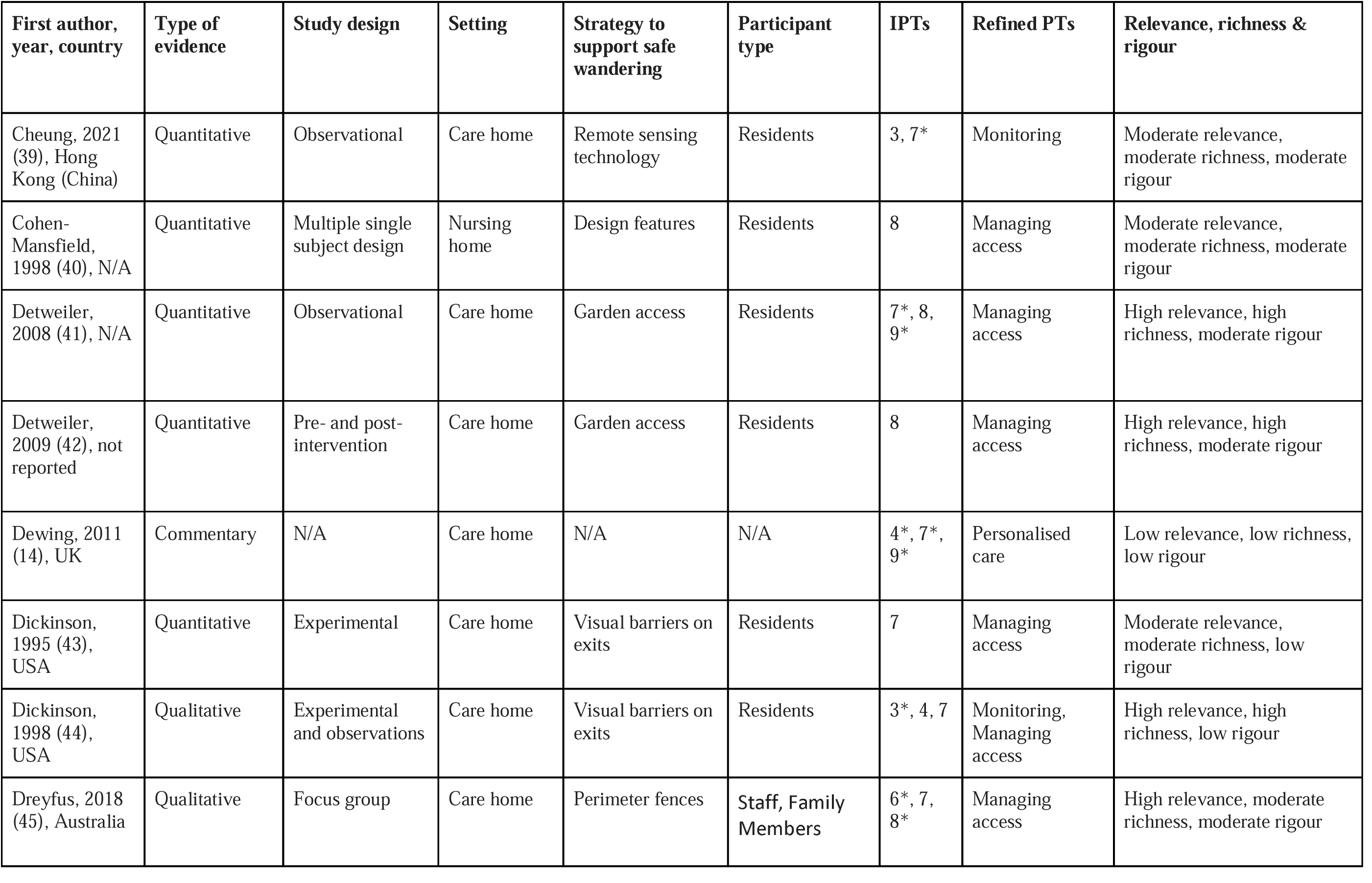
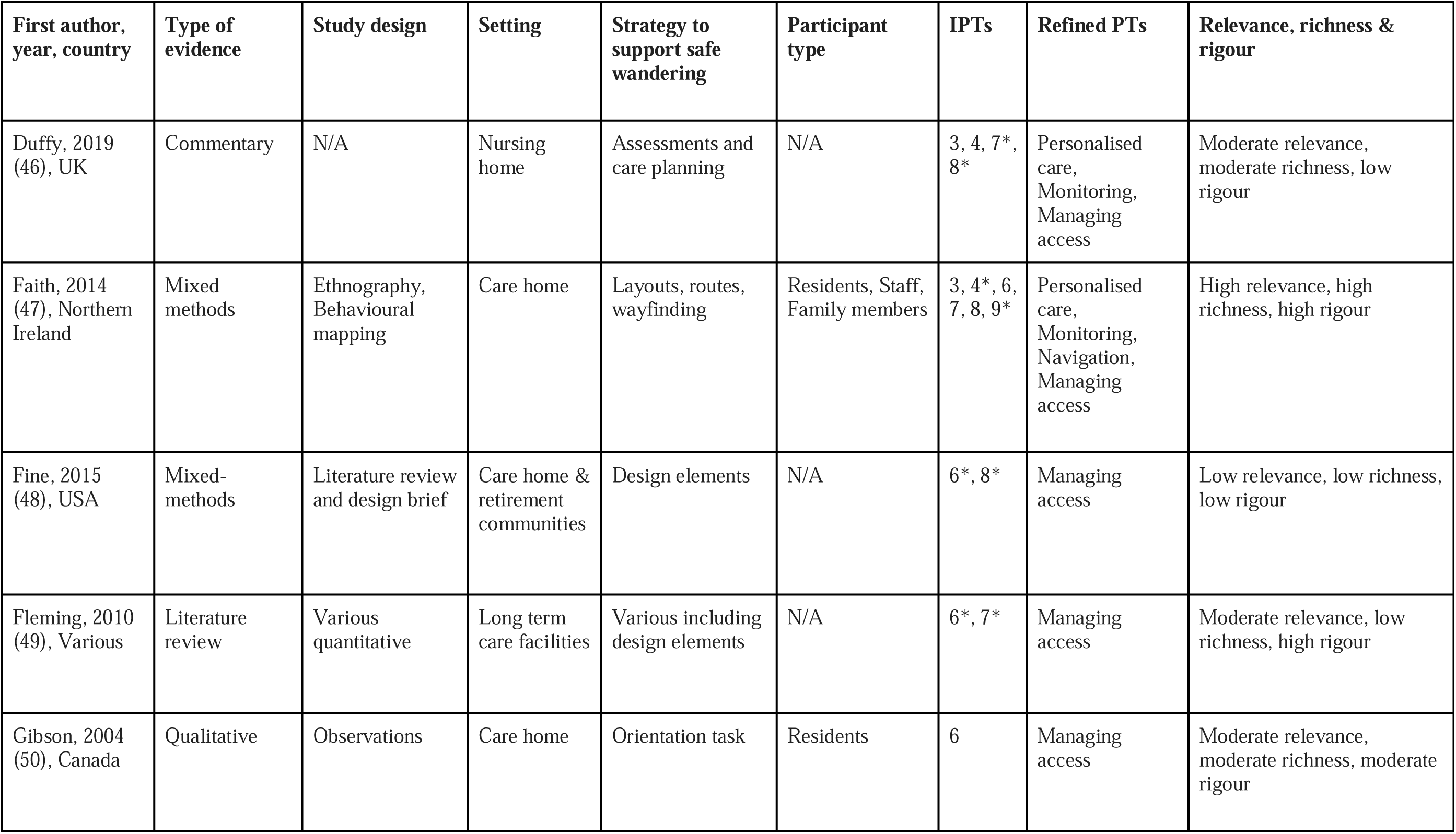
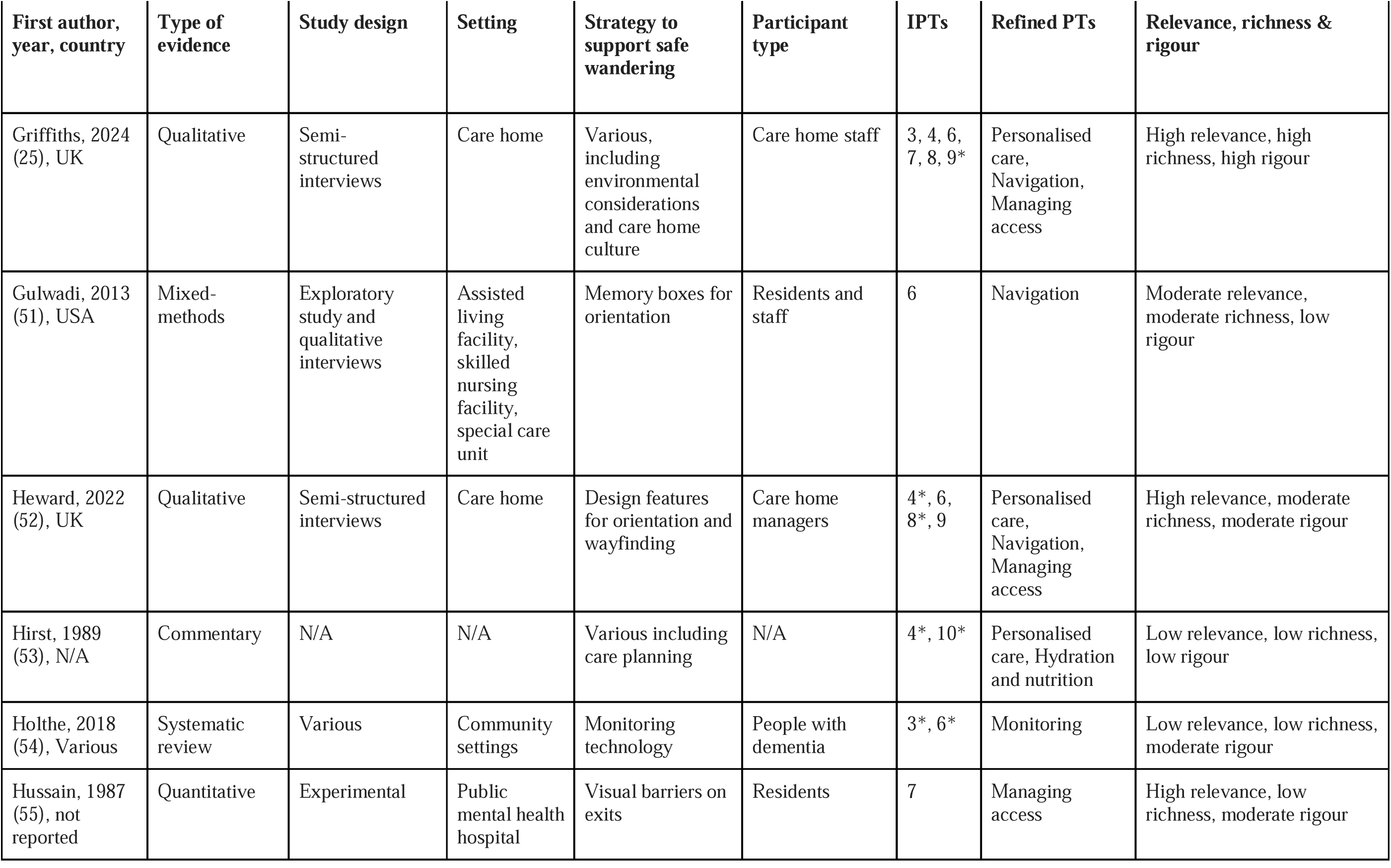
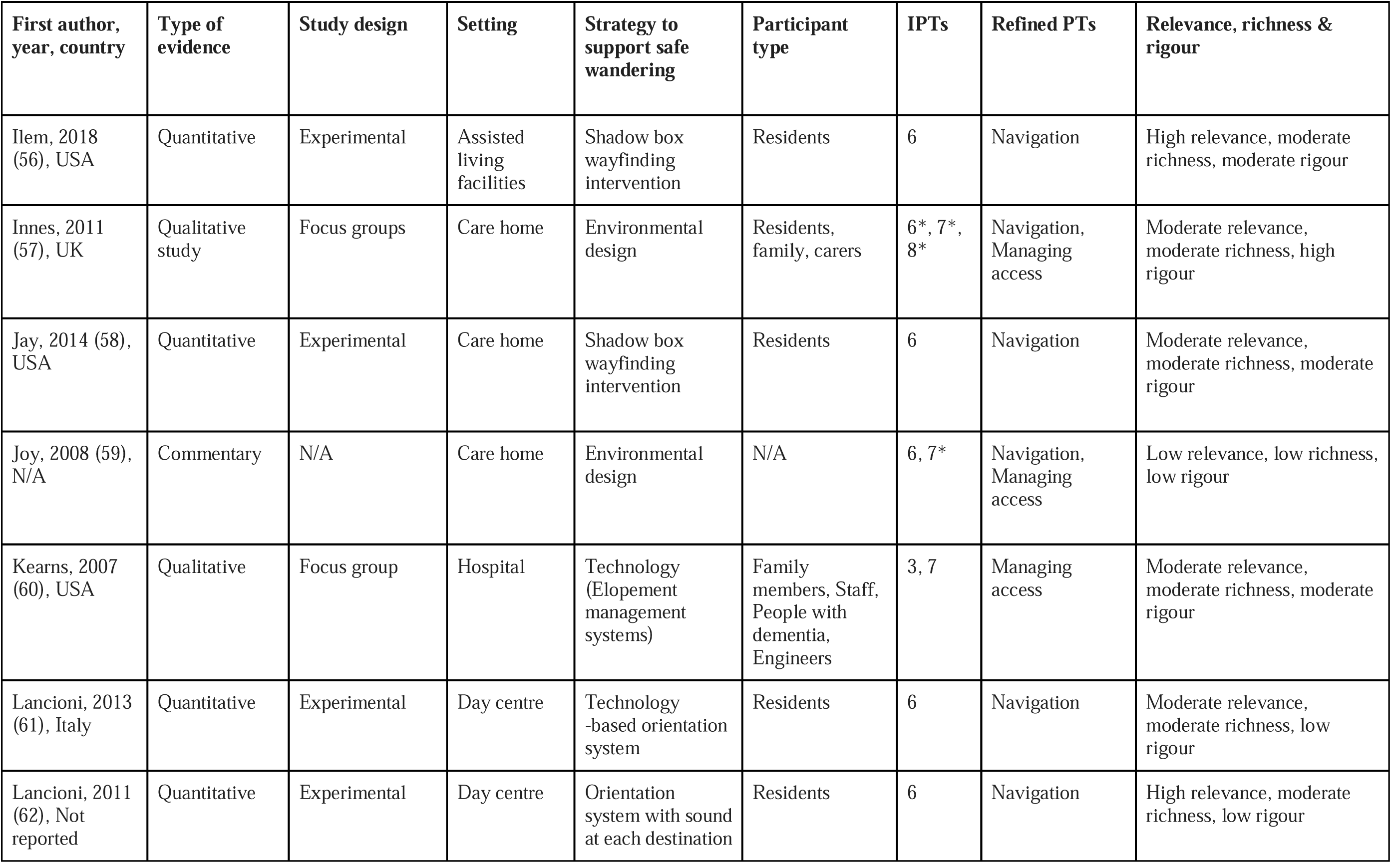
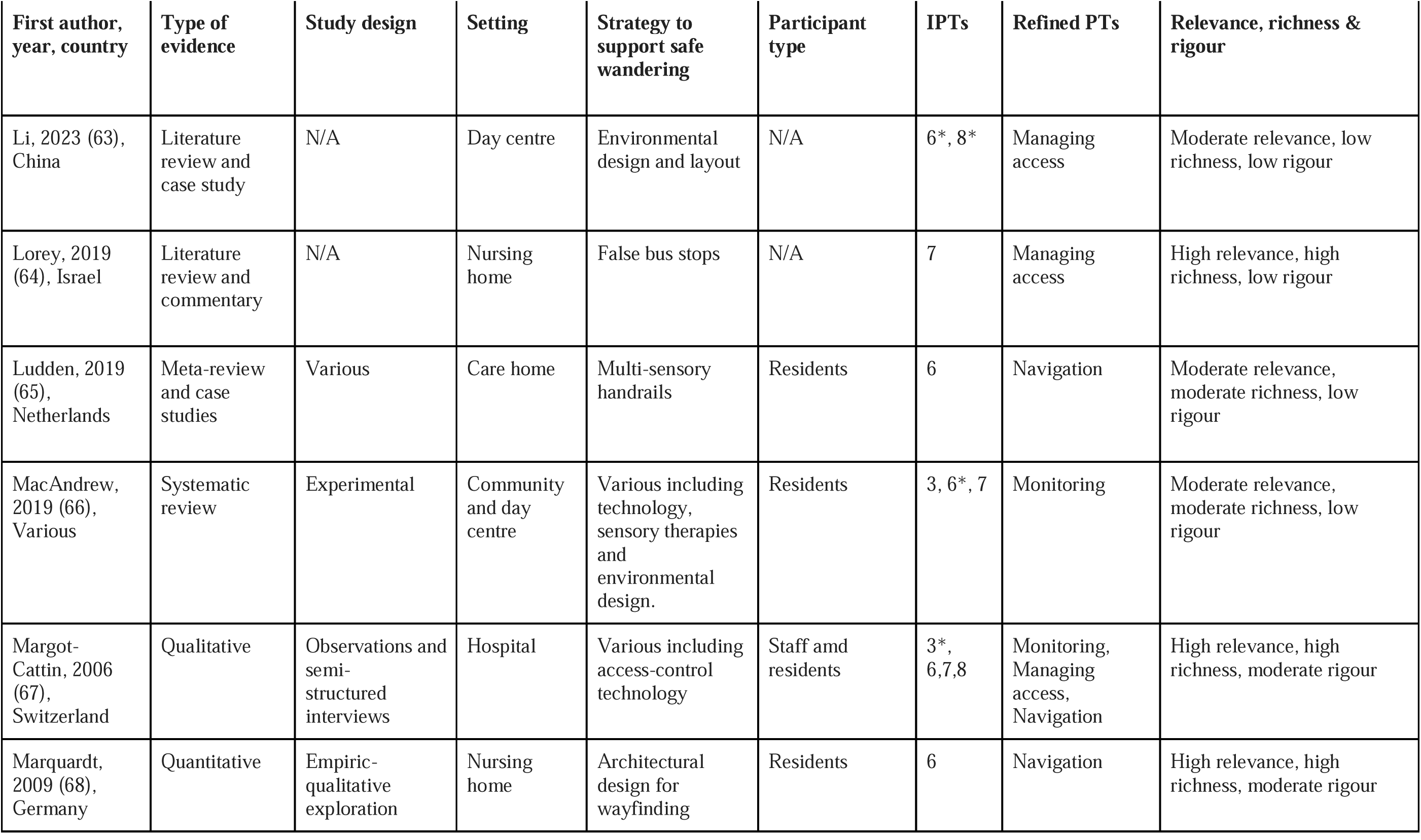
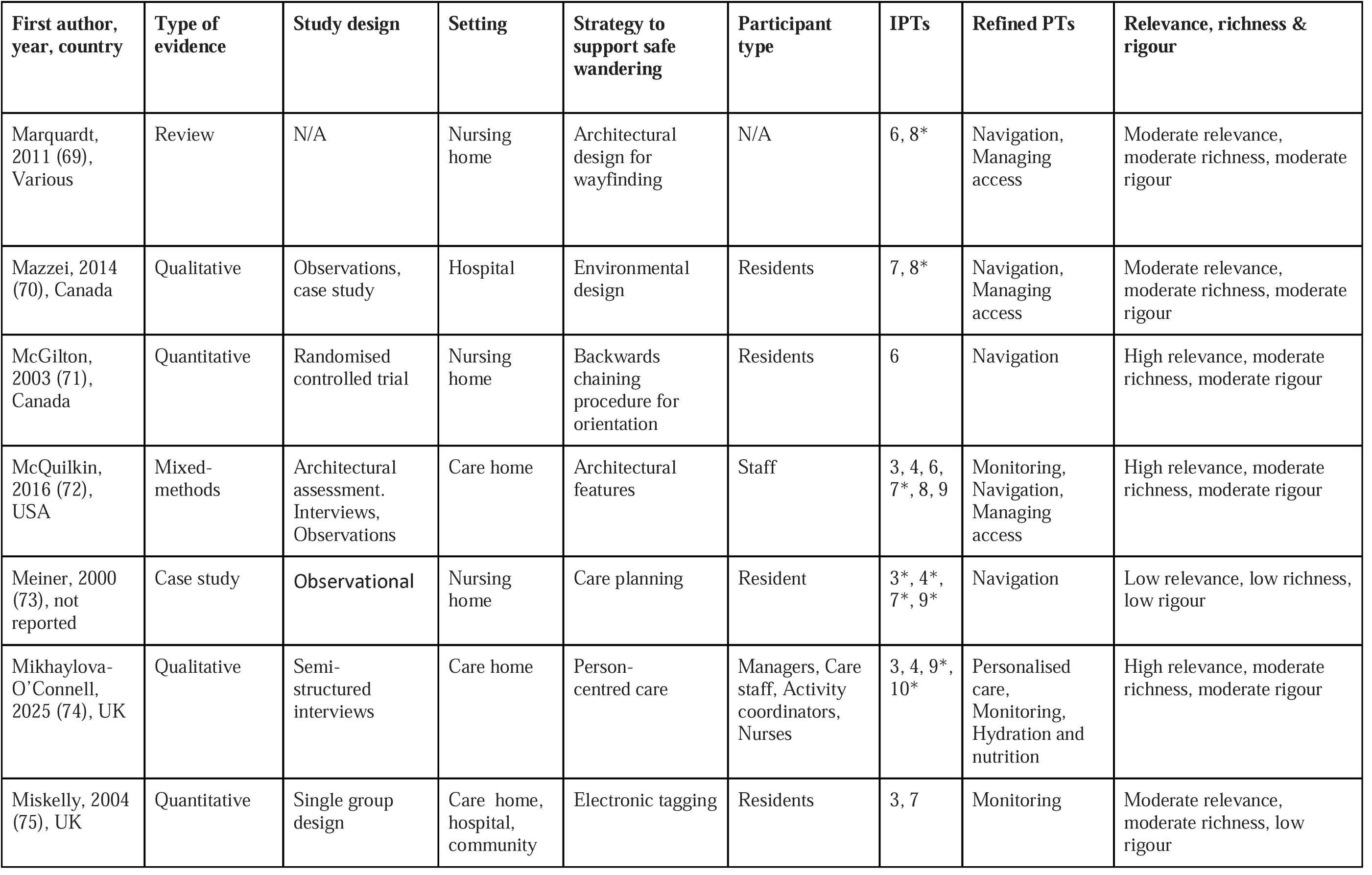
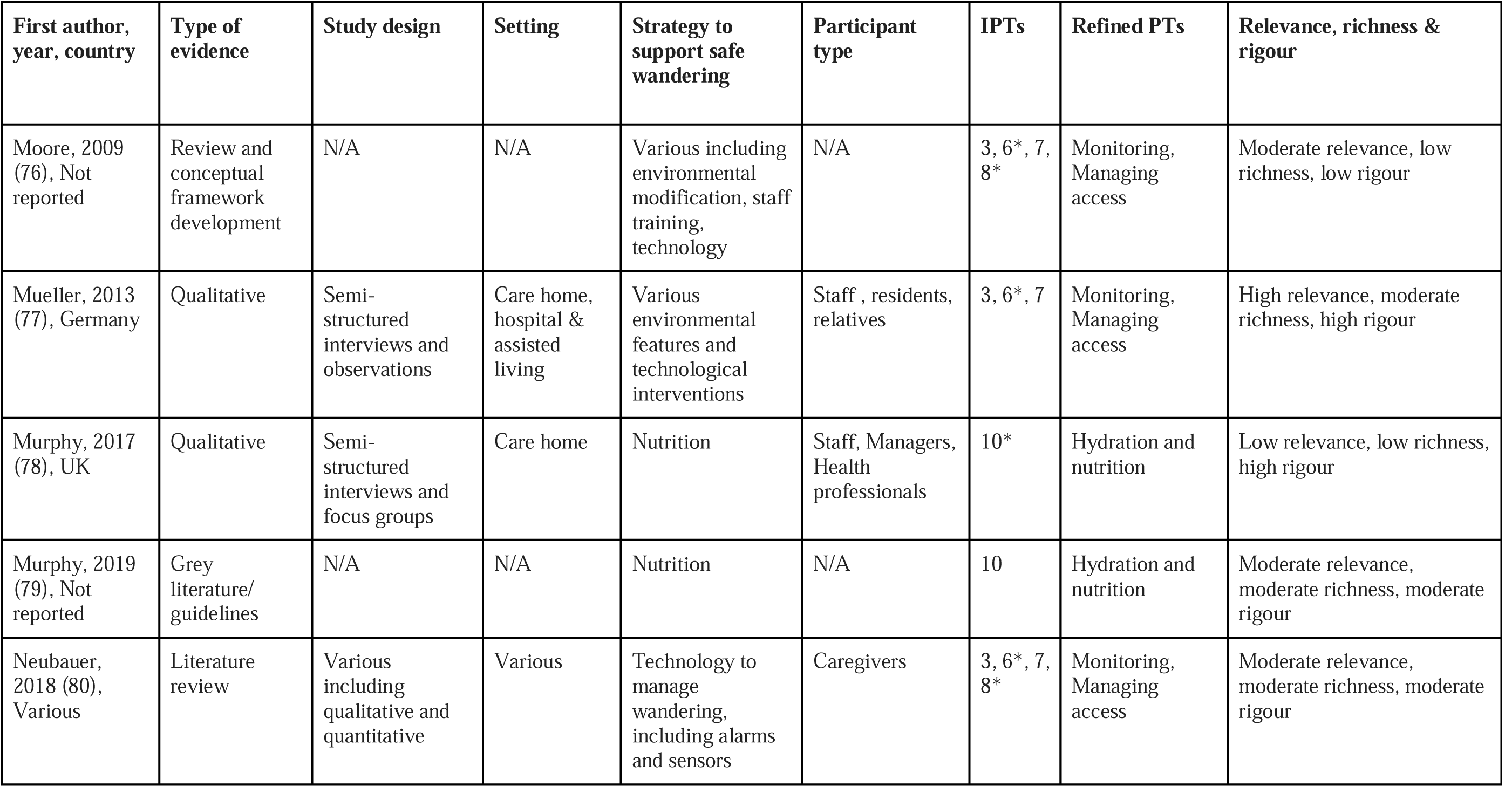
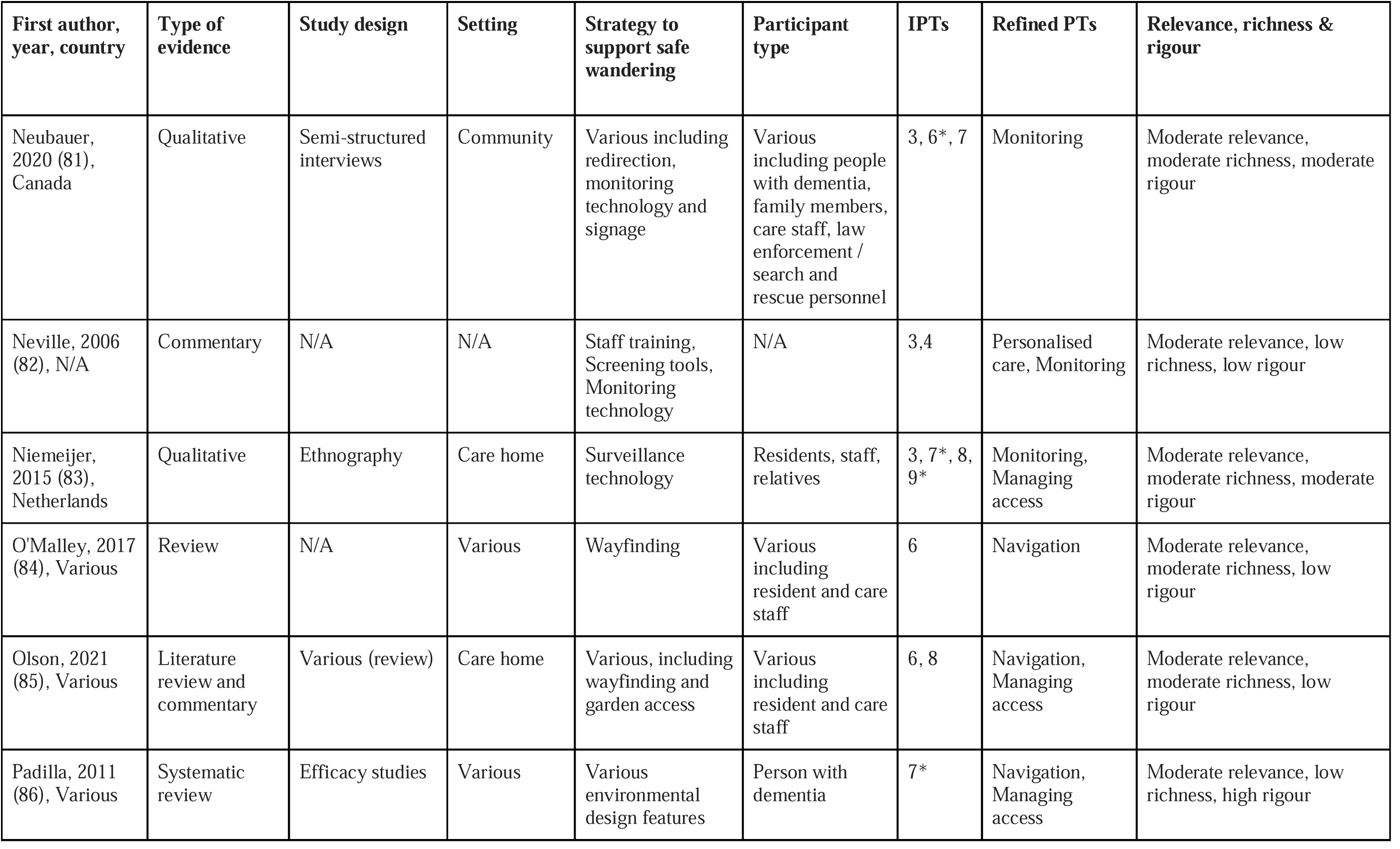
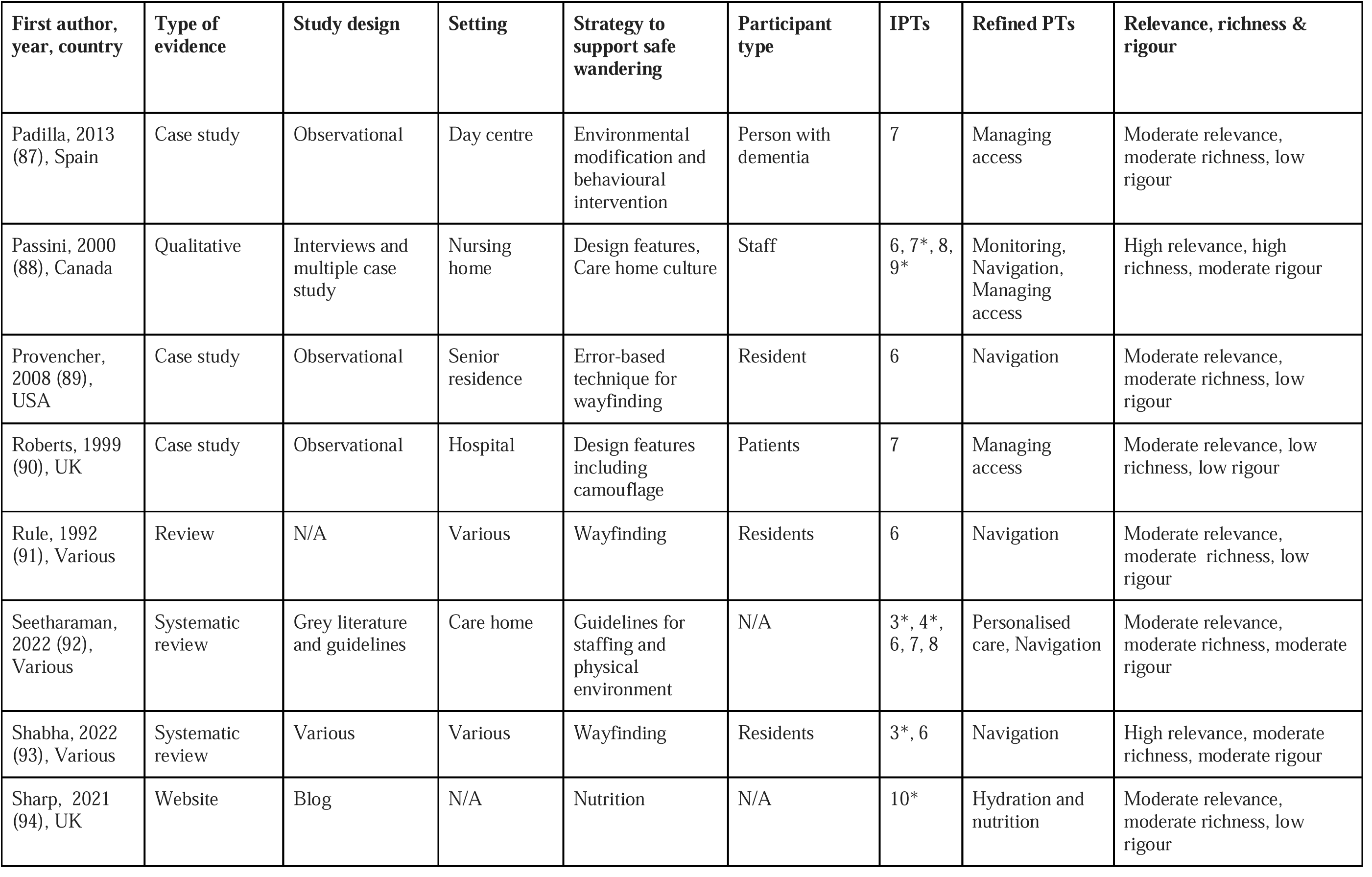
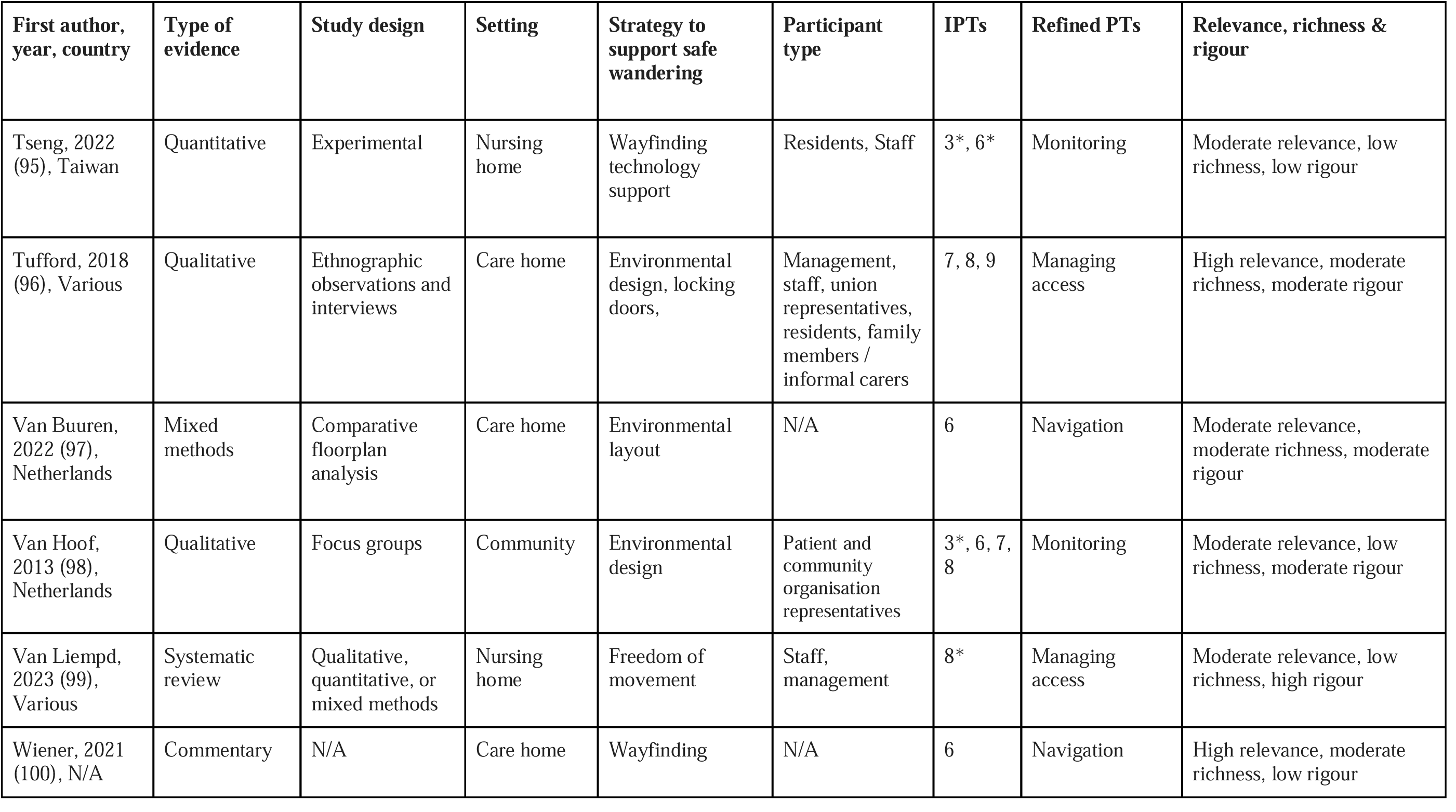
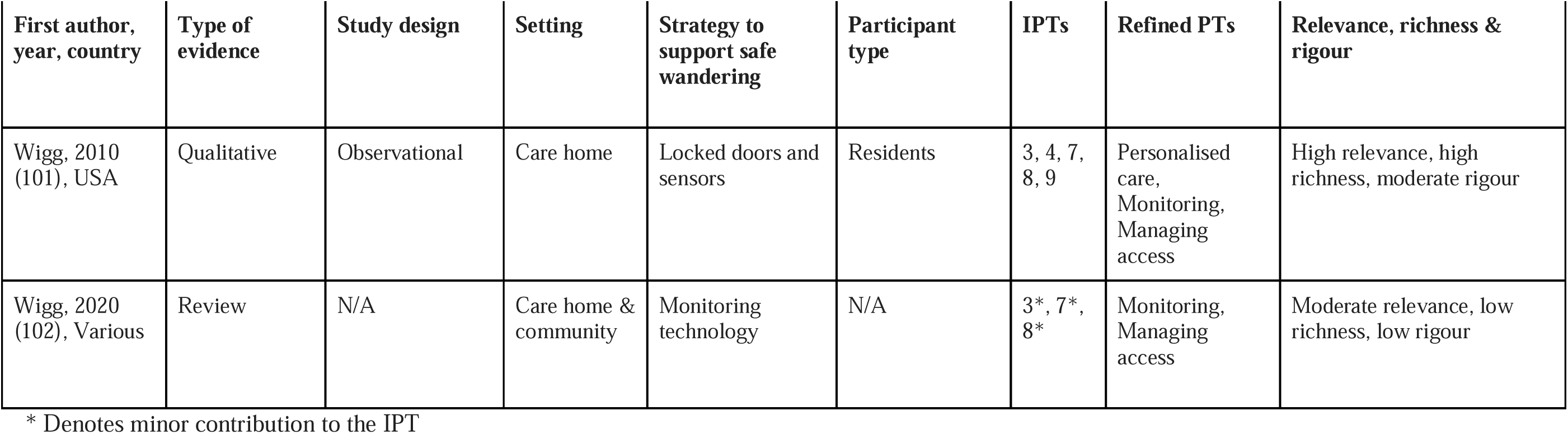
Characteristics of included studies.

| First author, year, country | Type of evidence | Study design | Setting | Strategy to support safe wandering | Participant type | IPTs | Refined PTs | Relevance, richness & rigour |
| --- | --- | --- | --- | --- | --- | --- | --- | --- |
| Alam, 2023 (28), USA | Mixed methods | Survey and interview | Care home | Design elements | Care staff | 3*, 6, 7* | Navigation | Moderate relevance, moderate richness, low rigour |
| Anderson, 2023 (29), Various | Review | Experimental | Assisted living | Environmental modification to prevent exit | Residents | 6*, 7, 8* | Managing access | High relevance, moderate richness, low rigour |
| Anonymous, 2007 (30), Not reported | Commentary | N/A | Care home | Various | N/A | 3*, 6*, 7* | Monitoring, Navigation, Managing access | Moderate relevance, low richness, low rigour |
| Anonymous, 2018 (31), UK | Website | Blog | Care home | Wayfinding intervention (signs) | N/A | 6* | Navigation | Low relevance, low richness, low rigour |
| Apple, 2015 (32), USA | Mixed-methods | Instrumental case study | Care home | Physical environment | Residents, Staff | 3,6,7,8, 9* | Monitoring, Managing access | Moderate relevance, moderate richness, moderate rigour |
| Barrett, 2020 (7), UK | Mixed-methods | Survey and interview, case studies | Extra care housing and retirement housing | Various, mainly focused on person-centred care | Managers, care staff and family members of residents | 3*, 4, 6*, 7, 8*, 9 | Personalised care, Monitoring, Managing access | High relevance, moderate richness, moderate rigour |

| **First author, year, country** | **Type of evidence** | **Study design** | **Setting** | **Strategy to support safe wandering** | **Participant type** | **IPTs** | **Refined PTs** | **Relevance, richness & rigour** |
| --- | --- | --- | --- | --- | --- | --- | --- | --- |
| Bautrant, 2019 (33), Not reported | Quantitative | Pre- and post-intervention | Care home | Environmental modification / Design elements | Residents | 6 | Navigation | Moderate relevance, low richness, low rigour |
| Benbow, 2017 (34), Various | Commentary | N/A | Care home | Environmental modification / Design elements | N/A | 3*, 6*, 7, 8 | Personalised care, Monitoring, Managing access | High relevance, moderate richness, moderate rigour |
| Bowes & Dawson, 2019 (35), Various | Systematic review | Various | Various including care homes | Environmental design features, Preventing exit | Various | 6, 7, 8* | Monitoring, Navigation Managing access | High relevance, moderate richness, high rigour |
| Britton, 2021 (36), UK | Website | Blog | N/A | Various including nutrition and environmental modification | N/A | 8*, 9*, 10* | Managing access, Hydration and nutrition | Moderate relevance, moderate richness, low rigour |
| Brush, 2008 (35), N/A | Commentary | N/A | Various | Wayfinding | NA | 6 | Navigation | High relevance, moderate richness, high rigour |
| Brush, 2015 (37), USA | Quantitative | Pre- and post-intervention | Care communities | Wayfinding intervention (signs) | Residents | 6 | Navigation | High relevance, moderate richness, moderate rigour |
| Caffo, 2014 (38), Italy | Quantitative | Experimental | Care home and day centre | Assistive technology / Orientation | Residents | 6 | Navigation | High relevance, moderate richness, moderate rigour |
| Cheung, 2021 (39), Hong Kong (China) | Quantitative | Observational | Care home | Remote sensing technology | Residents | 3, 7* | Monitoring | Moderate relevance, moderate richness, moderate rigour |
| Cohen-Mansfield, 1998 (40), N/A | Quantitative | Multiple single subject design | Nursing home | Design features | Residents | 8 | Managing access | Moderate relevance, moderate richness, moderate rigour |
| Detweiler, 2008 (41), N/A | Quantitative | Observational | Care home | Garden access | Residents | 7*, 8, 9* | Managing access | High relevance, high richness, moderate rigour |
| Detweiler, 2009 (42), not reported | Quantitative | Pre- and post-intervention | Care home | Garden access | Residents | 8 | Managing access | High relevance, high richness, moderate rigour |
| Dewing, 2011 (14), UK | Commentary | N/A | Care home | N/A | N/A | 4*, 7*, 9* | Personalised care | Low relevance, low richness, low rigour |
| Dickinson, 1995 (43), USA | Quantitative | Experimental | Care home | Visual barriers on exits | Residents | 7 | Managing access | Moderate relevance, moderate richness, low rigour |
| Dickinson, 1998 (44), USA | Qualitative | Experimental and observations | Care home | Visual barriers on exits | Residents | 3*, 4, 7 | Monitoring, Managing access | High relevance, high richness, low rigour |
| Dreyfus, 2018 (45), Australia | Qualitative | Focus group | Care home | Perimeter fences | Staff, Family Members | 6*, 7, 8* | Managing access | High relevance, moderate richness, moderate rigour |
| Duffy, 2019 (46), UK | Commentary | N/A | Nursing home | Assessments and care planning | N/A | 3, 4, 7*, 8* | Personalised care, Monitoring, Managing access | Moderate relevance, moderate richness, low rigour |
| Faith, 2014 (47), Northern Ireland | Mixed methods | Ethnography, Behavioural mapping | Care home | Layouts, routes, wayfinding | Residents, Staff, Family members | 3, 4*, 6, 7, 8, 9* | Personalised care, Monitoring, Navigation, Managing access | High relevance, high richness, high rigour |
| Fine, 2015 (48), USA | Mixed-methods | Literature review and design brief | Care home & retirement communities | Design elements | N/A | 6*, 8* | Managing access | Low relevance, low richness, low rigour |
| Fleming, 2010 (49), Various | Literature review | Various quantitative | Long term care facilities | Various including design elements | N/A | 6*, 7* | Managing access | Moderate relevance, low richness, high rigour |
| Gibson, 2004 (50), Canada | Qualitative | Observations | Care home | Orientation task | Residents | 6 | Managing access | Moderate relevance, moderate richness, moderate rigour |
| Griffiths, 2024 (25), UK | Qualitative | Semi-structured interviews | Care home | Various, including environmental considerations and care home culture | Care home staff | 3, 4, 6, 7, 8, 9* | Personalised care, Navigation, Managing access | High relevance, high richness, high rigour |
| Gulwadi, 2013 (51), USA | Mixed-methods | Exploratory study and qualitative interviews | Assisted living facility, skilled nursing facility, special care unit | Memory boxes for orientation | Residents and staff | 6 | Navigation | Moderate relevance, moderate richness, low rigour |
| Heward, 2022 (52), UK | Qualitative | Semi-structured interviews | Care home | Design features for orientation and wayfinding | Care home managers | 4*, 6, 8*, 9 | Personalised care, Navigation, Managing access | High relevance, moderate richness, moderate rigour |
| Hirst, 1989 (53), N/A | Commentary | N/A | N/A | Various including care planning | N/A | 4*, 10* | Personalised care, Hydration and nutrition | Low relevance, low richness, low rigour |
| Holthe, 2018 (54), Various | Systematic review | Various | Community settings | Monitoring technology | People with dementia | 3*, 6* | Monitoring | Low relevance, low richness, moderate rigour |
| Hussain, 1987 (55), not reported | Quantitative | Experimental | Public mental health hospital | Visual barriers on exits | Residents | 7 | Managing access | High relevance, low richness, moderate rigour |

| **First author, year, country** | **Type of evidence** | **Study design** | **Setting** | **Strategy to support safe wandering** | **Participant type** | **IPTs** | **Refined PTs** | **Relevance, richness & rigour** |
| --- | --- | --- | --- | --- | --- | --- | --- | --- |
| Ilem, 2018 (56), USA | Quantitative | Experimental | Assisted living facilities | Shadow box wayfinding intervention | Residents | 6 | Navigation | High relevance, moderate richness, moderate rigour |
| Innes, 2011 (57), UK | Qualitative study | Focus groups | Care home | Environmental design | Residents, family, carers | 6*, 7*, 8* | Navigation, Managing access | Moderate relevance, moderate richness, high rigour |
| Jay, 2014 (58), USA | Quantitative | Experimental | Care home | Shadow box wayfinding intervention | Residents | 6 | Navigation | Moderate relevance, moderate richness, moderate rigour |
| Joy, 2008 (59), N/A | Commentary | N/A | Care home | Environmental design | N/A | 6, 7* | Navigation, Managing access | Low relevance, low richness, low rigour |
| Kearns, 2007 (60), USA | Qualitative | Focus group | Hospital | Technology (Elopement management systems) | Family members, Staff, People with dementia, Engineers | 3, 7 | Managing access | Moderate relevance, moderate richness, moderate rigour |
| Lancioni, 2013 (61), Italy | Quantitative | Experimental | Day centre | Technology -based orientation system | Residents | 6 | Navigation | Moderate relevance, moderate richness, low rigour |
| Lancioni, 2011 (62), Not reported | Quantitative | Experimental | Day centre | Orientation system with sound at each destination | Residents | 6 | Navigation | High relevance, moderate richness, low rigour |
| Li, 2023 (63), China | Literature review and case study | N/A | Day centre | Environmental design and layout | N/A | 6*, 8* | Managing access | Moderate relevance, low richness, low rigour |
| Lorey, 2019 (64), Israel | Literature review and commentary | N/A | Nursing home | False bus stops | N/A | 7 | Managing access | High relevance, high richness, low rigour |
| Ludden, 2019 (65), Netherlands | Meta-review and case studies | Various | Care home | Multi-sensory handrails | Residents | 6 | Navigation | Moderate relevance, moderate richness, low rigour |
| MacAndrew, 2019 (66), Various | Systematic review | Experimental | Community and day centre | Various including technology, sensory therapies and environmental design. | Residents | 3, 6*, 7 | Monitoring | Moderate relevance, moderate richness, low rigour |
| Margot-Cattin, 2006 (67), Switzerland | Qualitative | Observations and semi-structured interviews | Hospital | Various including access-control technology | Staff and residents | 3*, 6,7,8 | Monitoring, Managing access, Navigation | High relevance, high richness, moderate rigour |
| Marquardt, 2009 (68), Germany | Quantitative | Empiric-qualitative exploration | Nursing home | Architectural design for wayfinding | Residents | 6 | Navigation | High relevance, high richness, moderate rigour |
| Marquardt, 2011 (69), Various | Review | N/A | Nursing home | Architectural design for wayfinding | N/A | 6, 8* | Navigation, Managing access | Moderate relevance, moderate richness, moderate rigour |
| Mazzei, 2014 (70), Canada | Qualitative | Observations, case study | Hospital | Environmental design | Residents | 7, 8* | Navigation, Managing access | Moderate relevance, moderate richness, moderate rigour |
| McGilton, 2003 (71), Canada | Quantitative | Randomised controlled trial | Nursing home | Backwards chaining procedure for orientation | Residents | 6 | Navigation | High relevance, moderate richness, moderate rigour |
| McQuilkin, 2016 (72), USA | Mixed-methods | Architectural assessment. Interviews, Observations | Care home | Architectural features | Staff | 3, 4, 6, 7*, 8, 9 | Monitoring, Navigation, Managing access | High relevance, moderate richness, moderate rigour |
| Meiner, 2000 (73), not reported | Case study | Observational | Nursing home | Care planning | Resident | 3*, 4*, 7*, 9* | Navigation | Low relevance, low richness, low rigour |
| Mikhaylova-O'Connell, 2025 (74), UK | Qualitative | Semi-structured interviews | Care home | Person-centred care | Managers, Care staff, Activity coordinators, Nurses | 3, 4, 9*, 10* | Personalised care, Monitoring, Hydration and nutrition | High relevance, moderate richness, moderate rigour |
| Miskelly, 2004 (75), UK | Quantitative | Single group design | Care home, hospital, community | Electronic tagging | Residents | 3, 7 | Monitoring | Moderate relevance, moderate richness, low rigour |
| Moore, 2009 (76), Not reported | Review and conceptual framework development | N/A | N/A | Various including environmental modification, staff training, technology | N/A | 3, 6*, 7, 8* | Monitoring, Managing access | Moderate relevance, low richness, low rigour |
| Mueller, 2013 (77), Germany | Qualitative | Semi-structured interviews and observations | Care home, hospital & assisted living | Various environmental features and technological interventions | Staff , residents, relatives | 3, 6*, 7 | Monitoring, Managing access | High relevance, moderate richness, high rigour |
| Murphy, 2017 (78), UK | Qualitative | Semi-structured interviews and focus groups | Care home | Nutrition | Staff, Managers, Health professionals | 10* | Hydration and nutrition | Low relevance, low richness, high rigour |
| Murphy, 2019 (79), Not reported | Grey literature/ guidelines | N/A | N/A | Nutrition | N/A | 10 | Hydration and nutrition | Moderate relevance, moderate richness, moderate rigour |
| Neubauer, 2018 (80), Various | Literature review | Various including qualitative and quantitative | Various | Technology to manage wandering, including alarms and sensors | Caregivers | 3, 6*, 7, 8* | Monitoring, Managing access | Moderate relevance, moderate richness, moderate rigour |
| Neubauer, 2020 (81), Canada | Qualitative | Semi-structured interviews | Community | Various including redirection, monitoring technology and signage | Various including people with dementia, family members, care staff, law enforcement / search and rescue personnel | 3, 6*, 7 | Monitoring | Moderate relevance, moderate richness, moderate rigour |
| Neville, 2006 (82), N/A | Commentary | N/A | N/A | Staff training, Screening tools, Monitoring technology | N/A | 3,4 | Personalised care, Monitoring | Moderate relevance, low richness, low rigour |
| Niemeijer, 2015 (83), Netherlands | Qualitative | Ethnography | Care home | Surveillance technology | Residents, staff, relatives | 3, 7*, 8, 9* | Monitoring, Managing access | Moderate relevance, moderate richness, moderate rigour |
| O'Malley, 2017 (84), Various | Review | N/A | Various | Wayfinding | Various including resident and care staff | 6 | Navigation | Moderate relevance, moderate richness, low rigour |
| Olson, 2021 (85), Various | Literature review and commentary | Various (review) | Care home | Various, including wayfinding and garden access | Various including resident and care staff | 6, 8 | Navigation, Managing access | Moderate relevance, moderate richness, low rigour |
| Padilla, 2011 (86), Various | Systematic review | Efficacy studies | Various | Various environmental design features | Person with dementia | 7* | Navigation, Managing access | Moderate relevance, low richness, high rigour |
| Padilla, 2013 (87), Spain | Case study | Observational | Day centre | Environmental modification and behavioural intervention | Person with dementia | 7 | Managing access | Moderate relevance, moderate richness, low rigour |
| Passini, 2000 (88), Canada | Qualitative | Interviews and multiple case study | Nursing home | Design features, Care home culture | Staff | 6, 7*, 8, 9* | Monitoring, Navigation, Managing access | High relevance, high richness, moderate rigour |
| Provencher, 2008 (89), USA | Case study | Observational | Senior residence | Error-based technique for wayfinding | Resident | 6 | Navigation | Moderate relevance, moderate richness, low rigour |
| Roberts, 1999 (90), UK | Case study | Observational | Hospital | Design features including camouflage | Patients | 7 | Managing access | Moderate relevance, low richness, low rigour |
| Rule, 1992 (91), Various | Review | N/A | Various | Wayfinding | Residents | 6 | Navigation | Moderate relevance, moderate richness, low rigour |
| Seetharaman, 2022 (92), Various | Systematic review | Grey literature and guidelines | Care home | Guidelines for staffing and physical environment | N/A | 3*, 4*, 6, 7, 8 | Personalised care, Navigation | Moderate relevance, moderate richness, moderate rigour |
| Shabha, 2022 (93), Various | Systematic review | Various | Various | Wayfinding | Residents | 3*, 6 | Navigation | High relevance, moderate richness, moderate rigour |
| Sharp, 2021 (94), UK | Website | Blog | N/A | Nutrition | N/A | 10* | Hydration and nutrition | Moderate relevance, moderate richness, low rigour |
| Tseng, 2022 (95), Taiwan | Quantitative | Experimental | Nursing home | Wayfinding technology support | Residents, Staff | 3*, 6* | Monitoring | Moderate relevance, low richness, low rigour |
| Tufford, 2018 (96), Various | Qualitative | Ethnographic observations and interviews | Care home | Environmental design, locking doors, | Management, staff, union representatives, residents, family members / informal carers | 7, 8, 9 | Managing access | High relevance, moderate richness, moderate rigour |
| Van Buuren, 2022 (97), Netherlands | Mixed methods | Comparative floorplan analysis | Care home | Environmental layout | N/A | 6 | Navigation | Moderate relevance, moderate richness, moderate rigour |
| Van Hoof, 2013 (98), Netherlands | Qualitative | Focus groups | Community | Environmental design | Patient and community organisation representatives | 3*, 6, 7, 8 | Monitoring | Moderate relevance, low richness, moderate rigour |
| Van Liempd, 2023 (99), Various | Systematic review | Qualitative, quantitative, or mixed methods | Nursing home | Freedom of movement | Staff, management | 8* | Managing access | Moderate relevance, low richness, high rigour |
| Wiener, 2021 (100), N/A | Commentary | N/A | Care home | Wayfinding | N/A | 6 | Navigation | High relevance, moderate richness, low rigour |
| Wigg, 2010 (101), USA | Qualitative | Observational | Care home | Locked doors and sensors | Residents | 3, 4, 7, 8, 9 | Personalised care, Monitoring, Managing access | High relevance, high richness, moderate rigour |
| Wigg, 2020 (102), Various | Review | N/A | Care home & community | Monitoring technology | N/A | 3*, 7*, 8* | Monitoring, Managing access | Moderate relevance, low richness, low rigour |

\* Denotes minor contribution to the IPT

### Realist synthesis

The overarching programme theory developed during the synthesis is presented in Figure 3. Within this overarching theory, we identified natural groupings within which distinct concepts could be arranged. These resembled the IPTs but were not exactly matching. We then grouped findings within these distinct concepts and synthesised them into the five refined programme theories resulting from natural grouping and distinct concepts identified within the overarching theory are: (1) personalised care; (2) monitoring; (3) navigation; (4) managing access; (5) hydration and nutrition (Table 2). Table S1 (Supplement 2) displays the evidence mapped to the IPTs and refined programme theories.

**Figure 3:**
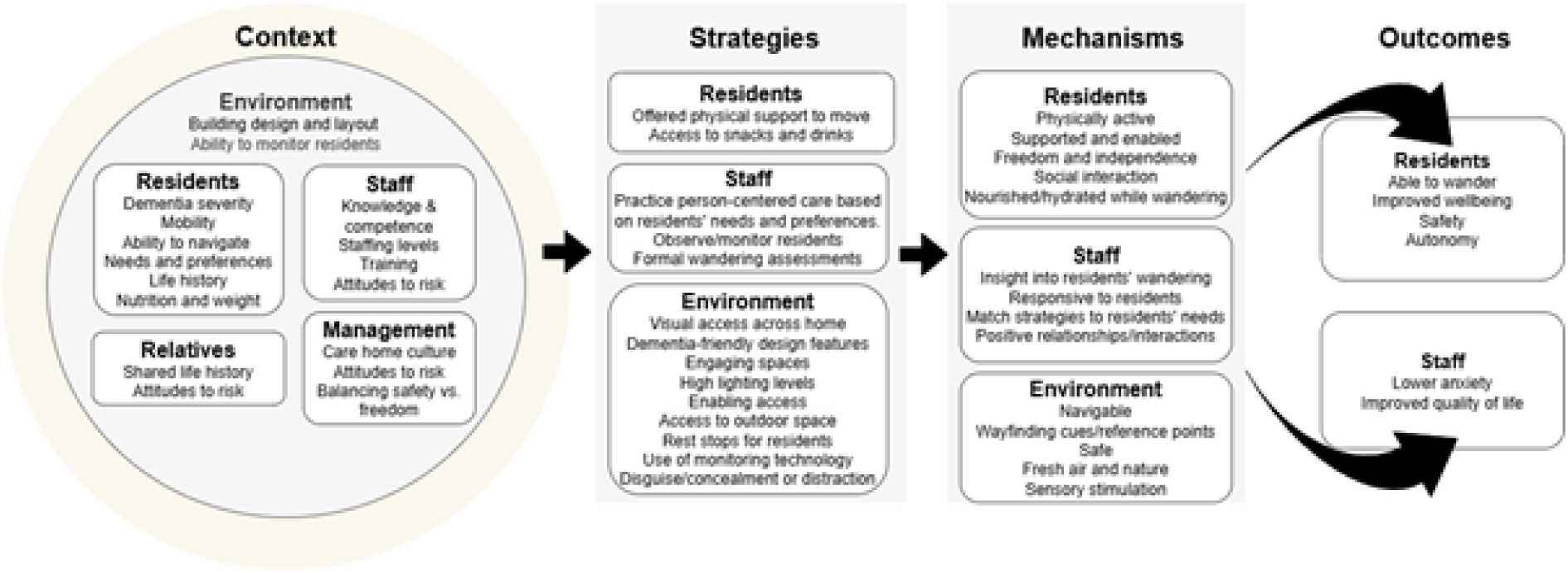
Overarching programme theory for safe wandering in care homes

**Table 2:** Refined programme theories.

| **Programme theory** | **Context** | **Strategy** | **Mechanism** | **Outcome** |
| --- | --- | --- | --- | --- |
| Personalised care | Staffing levels are suitable, with staff trained in person-centred care | Staff actively seek to know the residents and their histories, and practice person-centred care | Staff understand the reasons behind wandering, generally and in the moment, and match strategies to each resident. Staff and residents have positive relationships and interactions | Residents can wander, are kept safe, and have high well-being |
| Monitoring | Care home setup enables non-invasive monitoring, through good visual access or technological solutions | Staff monitor residents by keeping them in sight or with technological aids | Staff can detect when residents need support to walk, while also allowing residents freedom and independence | Residents experience autonomy resulting from independence, which improves their wellbeing. Residents can wander while staying safe from harm. Staff are less anxious. |
| Navigation | Design features are actively utilised to minimise the impact of residents' diminished orientation and wayfinding abilities due to dementia-related cognitive impairment | Dementia-friendly design features are incorporated into the environment to enable wayfinding. | The space becomes navigable even for residents unable mentally map their surroundings. Residents orient themselves and wayfind using environmental cues, and recognise familiar items and decor. | Residents have independence and autonomy in walking. Orientation and wayfinding difficulties |
| Managing access | Care home managers and staff must balance the need for resident safety with their need for freedom, independence and autonomy. This balance is informed by varying levels of risk aversion and risk-taking. | Residents have access to desirable/essential spaces, whilst being prevented from entering unsafe areas. This may involve locking/unlocking doors and using access control or alert technology. It may also involve strategies that leverage residents' visuospatial deficits to deter entry to particular areas. | Accessing spaces gives residents independence and a sense of autonomy, and restricting access to dangerous areas maintains safety. Access to outdoor spaces provides fresh air and a connection with nature. | Residents are kept safe from harm, and access and autonomy improve residents' wellbeing |
| Hydration and nutrition | Care homes seek to prevent residents who regularly wander from losing weight, and ensure residents are adequately nourished and hydrated even when they do not stay seated at mealtimes. Residents eat and drink as they walk. | Staff provide snacks and drinks in various locations, including on common wandering routes, ensuring availability of high-energy and nutritious food in small, accessible portions. | Residents can maintain their nutrition and hydration while walking as and when they wish, giving them a sense of autonomy. | Residents experience improved health and wellbeing. |

#### Personalised care

Within a person-centred care approach, wandering is seen as purposeful, and a culture of positivity about wandering and the freedom to take risks, supports staff in using individualised strategies to support residents to wander. Adopting a person-centred approach requires staff training and adequate staffing levels to provide personalised support, particularly for residents who wander.

##### Life history

Staff use strategies to understand each resident’s life history, gaining insights into the reasons why they are wandering. Ongoing discussions with relatives and documentation in care plans or the Alzheimer’s Society’s ‘This is me’ leaflet provide insight into residents’ previous careers, hobbies, and level of activity (7, 25, 52, 74, 101). Staff thus understand how walking and movement are integrated into each resident’s daily life, enabling provision of personalised and optimal support, including predicting which residents will wander (7, 14, 25, 34, 46, 52, 72). As a result, residents are supported in wandering, with improved mood and reduced anxiety and agitation (101).

##### Current motivations

Knowing each resident extends beyond life history knowledge to include interpersonal skills such as patience, reassurance, empathy and trust (52). Staff gain insights into residents’ experiences and motivations by speaking to them while walking, discussing what they are doing, and observing the way they interact with objects around the care home (34, 44, 74). Having sufficient staffing levels enables staff to spend time with residents to understand their reasons for wandering (7, 25, 74).

##### Formal wandering assessments

Formal wandering tools and assessments like the Dewing screening tool can be integrated into care protocols and highlight physiological or psychological needs that may precede wandering (14, 53, 82). Ongoing assessments and reviews ensure the use of person-centred strategies, allowing tailored care plans to be developed for each resident that support safe wandering (7).

##### Staff training

Staff interpersonal skills development is facilitated through training focused on supporting residents’ individual needs and enhanced through on-the-job learning, mentoring or shadowing (7, 25, 47, 92).

#### Monitoring

##### Visual monitoring

A layout with good visibility across the care home, including outdoor spaces, allows staff to monitor residents non-invasively (28, 47, 72, 101). Thus, staff can identify and assist residents who need help walking, enhancing their safety while promoting independence and autonomy (72). To improve monitoring, a smaller space with fewer residents and shorter corridors with no corners or kinks is ideal (32, 47, 98). Locating the staff base centrally can improve visual access (47, 88, 92) and larger homes can be separated into small clusters to facilitate visual monitoring (47).

##### Nighttime monitoring

Residents may be disorientated or confused when they get out of bed at night, and are likely to come to harm if they fall (47). Staff can keep residents safe through frequent monitoring (46), although poor staff adherence to checks may make this less successful (76).

##### Use of technology

Technological devices including cameras, sensors, door alarms, GPS trackers and motion detectors can enable staff to monitor residents by sounding an alarm or pushing a notification to their mobile device (7, 30, 39, 44, 54, 60, 66, 74–76, 80–83, 95, 98, 101, 102). Technology allows staff to monitor residents from a distance and be notified when assistance is needed (39, 80, 101), which affords residents independence and autonomy while maintaining safety (80). Residents have the freedom to access indoor and outdoor areas of the care home (80), while staff are reassured of residents’ safety, leading to reduced staff anxiety (95). This enables staff to engage positively with residents, leading to better interactions (101). Granting this freedom also reduces residents’ confusion, frustration and agitation when confronted with a physical barrier, such as a locked door (47, 99).

Technological failure, however, can lead to problems (67). Issues (e.g., an empty battery) may mean resident movement is not detected compromising residents’ safety (34, 77, 80, 83). Residents may remove a wearable monitoring tag if they feel stigmatised, uncomfortable with being monitored, or do not recognise it (35, 54, 67, 80, 81, 83). Conversely, staff may not respond to alerts, due to not noticing them, assuming someone else will respond, or irritation over frequent alerts (34, 83). Where technology is used to manage access, residents may experience distress upon witnessing another resident entering areas restricted to them (83). Staff may feel a (false) sense of security, leading to reduced attention to residents’ needs, and potentially management reducing staffing levels (67). Ethical concerns surround privacy and autonomy, with potential for monitoring to limit residents’ control, autonomy and freedom to wander (39, 47, 75, 83).

#### Navigation

##### Orientation and wayfinding difficulties

As dementia progresses, problems with orientation and wayfinding can worsen (68). Orientation is a resident’s sense of their place in the environment, while wayfinding is their ability to locate and move through spaces in the care home. As these abilities decline, a resident may find themselves in an unexpected place because of orientation difficulties, and familiar areas might seem out of place because of wayfinding difficulties. Residents may have sensory processing problems, visual/auditory impairments, and reduced ability to form and use a cognitive map, leading to deteriorating orientation (68, 69, 93). Cognitive maps are spatial representations of the environment that created in the mind’s eye, containing information on places beyond the range of vision (69). When disoriented, residents can experience helplessness, raised blood pressure, headaches, increased physical exertion and fatigue (103). Thus, orientation and wayfinding strategies are needed to help residents wander.

##### Wayfinding strategies

Strategies can be categorised into compensatory and restorative. Compensatory strategies provide residents with new ways to perform cognitive and behavioural tasks based on their existing competencies, while restorative strategies focus on restoring functioning (38). Common compensatory strategies include care home layout, colours and designs, signage, points of interest, personalised items, visual cues, and sensory stimulation. Restorative strategies include wayfinding training (35, 38, 47, 61, 62). Strategies are more effective when care quality is high, and when staff implement person-centred care, tailoring strategies to each resident (35) (see ‘Personalised care’).

##### Care home layout

Simple, compact care home layouts with good visual access aid navigation when residents can see their destination (68, 69, 84, 97). Walking around corners, decision-making at junctions and navigating dead-end corridors make wayfinding difficult, as the destination is not visible (68, 84, 97) and decision points can cause confusion (47). A home-like layout can aid navigation (97). Placing a kitchen or dining room at points where the direction changes can aid navigation as a meaningful anchor point (69).

Circular layouts support wandering by allowing residents to walk continuously without reaching dead ends (47, 70), while square-shaped layouts can be challenging due to frequent direction changes (47, 68, 69). Whether square or circular, continuous layouts facilitate wayfinding by allowing residents to walk without the need to orient themselves (47, 88). However, this can also lead to fatigue and exhaustion, especially in residents who tend to walk continuously (47).

A symmetrical layout can aid navigation when the two sides are sufficiently distinct, aiding development of a mental map (100). When both sides look identical, residents experience confusion and struggle with orientation and wayfinding, as spaces are not in expected locations (28, 47). Open-plan layouts can aid navigation by providing visual access to destinations (72, 93), however residents may struggle to distinguish between spaces unless they are well-defined, presenting a navigational challenge (47). In non-open-plan layouts, visual access can be improved by adding glazing and large doorways, giving residents sight of their destinations (47, 57, 67, 72). Placing spaces closer together and ensuring a clear path through the care home can enable wayfinding (25, 47, 92). Wide corridors allow residents ample space to wander, enhancing residents’ sense of freedom (25, 97).

##### Points of interest

Care home layouts can facilitate navigation and orientation by making certain spaces prominent, and therefore more meaningful and memorable to residents. A well-stocked, live-in kitchen can act as a focal point, particularly if there is only one available, or if multiple kitchens are positioned side-by-side (68, 69). Regardless of layout, navigation can be aided by visual cues and points of interest. Distinct spaces, in both function and design, that are architecturally legible, improve navigation (47, 68, 69, 84). Residents can recognise spaces more easily based on their function, aiding orientation (47, 52, 68, 69, 92). Rooms with multiple functions (e.g., dining room also used for activities), can cause confusion (47, 92).

Points of interest located throughout the care home (e.g., murals, paintings, clocks, furniture, memory boxes, architectural elements, the lift) can function as landmarks, aiding wayfinding (28, 51, 56, 58, 59, 68, 72, 84, 100), especially when hallways are similar (30). Landmarks help residents recognise their position and orientate towards their destination (30, 91, 100), particularly if they are unique, salient, distinctive and persistent (37, 68, 84, 100), and easy to recognise (100). ‘Beacons’ (landmarks viewable from decision points) are particularly effective for navigation (84, 100), especially when they are nameable (e.g., clock) (100). Points of interest encourage purposeful movement, attract residents’ attention to certain areas and start conversations (e.g., shop front displays) (25, 47). Adding furniture along long corridors for rest stops can alleviate tiredness and encourage walking (25). Some points of interest, however, can be confusing for instance, residents trying to interact with a mural (25). Similarly, residents can become disorientated and confused when furniture arrangements change unexpectedly, leading to falls (28).

##### Colour and design

Colour and design can aid orientation and wayfinding. Consistent use of colours throughout the care home helps residents interpret their environment (37) and varying colours by location enables residents to orient themselves (28, 52, 57, 91). Painting areas and features differing colours (e.g., residents’ room doors) can distinguish the spaces, aiding navigation (28, 52). Creating colour contrast between different elements (e.g., handrails and walls), can assist residents with visual impairments to perceive and identify key features, improving wayfinding and reducing falls (37, 47, 85). Since residents often look downwards when walking, floor coverings varying in colour can facilitate wayfinding (28, 47). Plain floor coverings without patterns or swirls can be easier for walking, as residents may perceive patterns as holes or water due to dementia-related visuospatial distortions (28, 47, 69, 85). Handrails provide a guide for residents to follow with stopping points when they end and support walking for longer distances (47). Bright lighting (natural and artificial) ensures residents can see well, even with visual impairments, and enables detection of wayfinding cues (33, 47, 97, 103).

##### Signage

Signs that are easy to see, understand and follow guide residents, aiding navigation (25). Low positioning (including on the floor), aligns with residents’ downward gaze, making signs easier to see (28, 30, 37, 47, 69, 92, 100, 103). Placing signs in uncluttered areas reduces the chance of information overload (69), and simple, uncluttered signs with large text, sans serif fonts, bold colours, and high colour contrast enhances visibility and clarity (37, 58). Signs combining words with pictograms are easier to follow and do not rely on word recognition (37, 47, 52, 69), particularly when using representational images (100). Naming and labelling areas of the care home (e.g., door numbers, ‘street’ addresses or name labels for residents’ rooms) can further aid navigation (28, 47, 51, 86), especially when residents can recall their door number or ‘address’, perhaps if written on their key (31, 47). However, a room number that differs from a resident’s previous house number can inhibit recall, as past addresses may interfere with learning the new one (47).

##### Multi-sensory stimulation

Multi-sensory stimulation can aid wayfinding by providing multiple types of information (28, 69, 103). If one sense is impaired, residents can use another (69), and integrating stimuli from various senses and sources creates a coherent environment (37, 65). Sensory cues aid orientation when coherent with the space (e.g., TV sounds in the living room, coffee and food smells from the kitchen/dining area) (28, 47). However, overbearing sensory stimulation (e.g., loud sounds) can cause distress.

##### Support from staff

Staff can support wayfinding through physical assistance, helping residents move in the right direction while also providing social support and interaction, benefiting residents’ well-being (14, 47, 52, 73, 83, 91). However, frequent accompaniment can reduce wayfinding abilities, potentially reducing autonomy, by giving residents fewer opportunities to practice their skills (88).

##### Route learning

Restorative strategies aiming to improve route learning can be successful when the surroundings are facilitative, for instance, a simple floor plan and key spaces located in close proximity (38). Multiple trials in relatively quick succession are required for successful route learning, but some residents may forget the route once training ends (71, 89). Access control systems (e.g., keycard access to bedrooms) support wayfinding by providing reinforcement, assisting recognition of residents’ own rooms and reducing the likelihood of unsuccessful door-testing (67).

#### Managing access

##### Balancing safety and freedom

Concerns about residents’ safety from management, care staff and relatives provide an overarching context for preventing residents from entering unsafe spaces. Different levels of risk aversion influence the strategies implemented, creating tension between safety and freedom for residents (25). Additionally, care home managers may worry about potential legal consequences of if residents come to harm (101).

##### Access to spaces within the care home

Access can be facilitated by a simple, compact, open-plan layout (see ‘Navigation’) situated on a single floor, with a wandering path throughout the home, and clearly visible common spaces located within close proximity (29, 34, 47, 63, 88, 92). Unlocked doors throughout the care home can reduce anxiety and improve sleep in residents (47, 83, 96, 99, 101).

Access to spaces throughout the care home is enhanced and enabled by ensuring that fixtures and fittings are obvious and easy to use. Door handles increase accessibility over door knobs (47), and bold colours and colour contrasts can make handrails, door features and spaces easy to see and navigate (47, 50, 52, 92) (see ‘Navigation’). Visually enhancements can enable access by making spaces appealing, for instance, by depicting natural scenery (40). Maintaining visual access throughout the care home and to outdoor areas increases light levels and promotes freedom of movement (47, 48, 72). Good lighting enables access to spaces, by compensating for age-related visual impairments (34, 47). Access is further enabled by floor coverings that are plain and simple, without any patterns that could be interpreted as holes, steps, water or obstructions (32, 47, 88, 92, 93). Hard floors are easier for residents to walk on than carpet, whereas uneven flooring and steps can increase falls (36, 47).

##### Access to outdoor space

Granting residents access to indoor and outdoor spaces in the care home can improve wellbeing and a sense of freedom and autonomy (25, 47, 57, 72, 101). Access can reduce agitation, frustration and distress, improve mood, quality of life, sleep quality, memory, language abilities, spatial abilities and energy levels and allow residents to escape excessive stimulation, noise and crowding inside the care home (25, 42, 47, 72, 80, 85, 96, 99, 101), in addition to improving the quality of life of staff members correspondingly, as residents are less agitated (41). Outdoor spaces can benefit residents through physical activity, exposure to fresh air, sunlight and greenery, contact with plants and animals, and exposure to seasonal change (47, 69, 72, 99, 101), in addition to increased and varied opportunities for social interaction with relatives and care staff (41, 57). Outdoor access is facilitated by ensuring such spaces contain shelter, adequate lighting, year-round access, ramps, handrails, sufficient interest (including places with unique character and intense activity), rest stops/seating, and wide walking paths with safe, level surfaces (29, 32, 47, 48, 57, 69, 72, 76, 102). Staff can facilitate and support access to outdoor spaces by monitoring residents (see ‘Monitoring’) and ensuring they have appropriate clothing for the weather (72). Doors leading outside from the live-in kitchen and/or residents’ bedrooms facilitate access to outdoor spaces (48, 69, 92, 96).

##### Preventing access to unsafe spaces

Several strategies have been employed by care home staff and management to prevent residents from accessing places that may present a safety hazard, to allow them to wander safely in other areas.

##### Camouflage or illusion

Camouflage or illusion can be used to facilitate wandering in certain areas of the care home. This included camouflaging doors to unsafe spaces (28, 29, 34, 35), such as disguising them as a continuation of the wall or with a mural, or disguising distinguishing features on a door such as the panic bar, door handle, using frosted glass, placing a mirror on the door, or placing markings in front of the door (e.g., tape placed across the floor as a two-dimensional grid) (7, 28, 29, 34, 43, 44, 49, 55, 59, 77, 86, 87, 90). Camouflage is most effective when lighting is good and the level of contrast is not too high or low (29). Residents often avoid camouflaged exit doors because visual and spatial perception issues prevent them from recognising the door (28, 34, 59). When residents move away from a disguised door on their own, staff spend less time redirecting them and, therefore, have time for positive interactions (29).

Residents with less significant cognitive impairments may still recognise a camouflaged door, leading to an increase in door-testing (29, 44, 70). Placing a mirror in front of a door can cause symptoms associated with agitation in some residents (86).

Disguising spaces poses potential ethical challenges due to inherent deception, although it could be argued that if the act of deception improves the well-being of residents it may be warranted (64). Residents who cannot perceive the exit door may find exiting the care home challenging in a fire (30), although staff support and additional signage can mitigate this risk (34). Positioning doors to limit visual access from communal and circulation spaces can reduce residents’ exiting because due to the lack of a visual exit cue (29, 34, 47). Door positioning can also reduce residents’ distress and behaviour that challenges by preventing them from feeling excluded from spaces (34, 47). This approach may circumvent some of the difficulties of camouflage and concealment approaches where the layout permits.

##### Locking doors

Keeping doors to exits and other unsafe spaces locked provides a physical barrier, preventing residents from coming to harm and therefore allowing them to wander safely in other areas)(7, 34, 70, 101). Locked doors are typically found in care homes with a culture of perceiving wandering as risky and risk as unacceptable, meaning individuals are prevented from independently accessing exits (101). When residents move through or towards a locked door (e.g., by following another resident and/or their family outside), staff redirect the resident (101). Residents can feel confined when outdoor spaces are fully enclosed with fences, and, in some cases, attempt to climb the fence (45). However, when residents experience freedom of movement in outdoor spaces, fences can benefit residents’ wellbeing as well as providing safety (7, 45, 101), and can create a sense of ownership over the space by defining boundaries, like front and back gardens at home (45). Residents living in settings with secure perimeter fences and unlocked doors are outdoors more often and are more physically active, leading to a better quality of life (45, 99, 101).

##### Use of technology

Wearable technology can control access by granting residents entry to safe spaces while restricting access to unsafe/private spaces (30, 46, 60, 67, 77). These devices can improve residents’ wellbeing by enabling them to walk freely without feeling confined (67). Residents experience increased security and improved self-esteem due to increased autonomy (67), although there are some potential issues with device use (see ‘Monitoring’).

##### Use of distraction

Distraction or stimulation, such as visual or tactile interest, art-based design or physical guidance from staff, can keep residents away from unsafe spaces by engaging their interest in the environment (29, 34, 44, 72, 87). Creating a circular ‘endless’ hallway can draw residents away from unsafe spaces by enabling them to walk continuously without reaching an exit (77). Creating an association between an unsafe space and a noxious stimulus can prevent residents from approaching that space (29).

#### Hydration and nutrition

Staff support the nourishment and hydration of residents as they wander by making food and drink more convenient and regularly available (53, 78). Food and drink can be placed along wandering routes or at endpoints, such as the living room (74, 79). Staff can also provide food and drink that residents can eat while on the move, including lunchboxes with separate sections, finger foods and grazing menus, and meal shakes to drink on the move (74, 78, 79, 94). Residents can thus eat and drink frequently, at their own pace while also wandering, enabling them to maintain nutrition and hydration (74, 78, 79, 94). This is particularly important for residents who do not stay seated at mealtimes (79, 94). Staff can increase residents’ nourishment by offering food high in energy and protein, providing energy to wander while preventing weight loss (79).

##### Personalised care

Knowing residents’ wandering patterns enable staff to provide food and drink according to residents’ preferences and patterns, supporting residents’ nutrition and hydration (74, 78, 79, 94). Monitoring residents’ weight can enable staff intervention to prevent (further) weight loss as part of delivering personalised care (74).

##### Safety considerations

Ensuring safe eating practices can facilitate safe wandering. Staff can support residents to eat and drink safely on the move by using appropriate lids on drinks bottles and ensuring finger foods are cut up small enough to be chewed and swallowed safely (36).

### Sense-checking conversations

To assess the relevance of review findings to current practice, 16 care home staff from 5 care homes participated in sense-checking focus groups (n = 13) and one-to-one interviews (n = 3). Participants were regional managers, team leaders, nurses, carers, front-of-house managers, and housekeeping staff.

Across the homes, staff expressed positive views about wandering, wishing to encourage this when possible, as it promotes physical activity and allows residents to achieve tasks or find purpose through household activities. Staff felt that preventing or dismissing wandering could increase falls, agitation, and behavioural issues among residents.

Low staffing levels limit visual monitoring and hinder staff’s ability to meet residents’ broader needs. Understaffed teams are more likely to discourage wandering and encourage residents to stay seated or remain in one room. One care home wanted to provide one-to-one support to residents who wander, but local authorities assessed this level of care unnecessary.

Care homes emphasised that monitoring should be a shared responsibility among all staff, not just carers. Non-care staff may have the time to spend with residents engaging in non-care tasks, and are generally freer to move around with residents. However, they cannot access residents’ care plans, limiting their provision of person-centred care. Additionally, staff reported challenges when supporting residents physically, particularly shorter carers who face increased risk of injury to themselves.

Staff members reported facing emotional challenges, as they feel mentally taxed when residents become agitated and distressed when they fall. Staff feel overwhelmed when multiple residents are walking simultaneously, underscoring the need for person-centred strategies.

#### Personalised Care

All participants emphasised the importance of understanding residents’ life histories. However, staff at one home reflected that knowing a resident’s background helps to contextualise their actions, but does not necessarily assist in supporting their wandering behaviour. New staff members may struggle to get to know residents, often relying on more experienced colleagues and relatives for additional information. One individual highlighted that when residents moved from hospitals, planning was compromised due to poor information on mobility from the hospital.

#### Monitoring

For visual monitoring, staff suggested asking residents to sit down when they appear unsteady, while watching them from a safe distance. The main devices used were room alarms and pressure mats or cushions. Many staff were unfamiliar with other devices discussed in the literature. One home used a nighttime audio listening device. In one home, management tracked staff response times to call alerts, which encouraged timely attendance, a common issue in homes without such oversight. They also mentioned using monitoring devices to track and review residents’ activity, food and drink intake. However, care homes highlighted issues such as equipment damage by residents and safety concerns related to spills. Some residents learned to bypass the devices, such as stepping over mats, which can increase risks.

#### Navigation

Navigation strategies resonated with staff, including signage, pictures on bedroom doors, coloured doors, and murals. However, some questioned the effectiveness of these for residents with advanced dementia, reflecting that they physically guide residents when they appear lost, which again highlights staffing concerns. One care home noted that communication difficulties with staff from other countries complicates navigation.

#### Managing Access

Staff felt it would be unfair for some residents to have access to certain areas while others do not. Some staff endorsed using distractions, and optical illusions to prevent residents from entering certain places. While some staff did not advocate for restricting access to certain areas, they acknowledged that some spaces should be locked for safety reasons (e.g. kitchens, cleaning cupboards, sloshies). While many care homes did not offer free garden access, staff said residents could ask to go outside. However, it was unclear whether residents had the capacity to understand this or make such requests.

Participants highlighted the importance of preventing residents from entering each other’s rooms, and how staff need to consider how the behaviour of one resident may affect others.

#### Hydration and Nutrition

Whilst in one care home, concerns about leaving snacks out existed for medical reasons, in another, snack trolleys and drinks were available at every seating location. Staff reported initial concerns about residents spilling liquids, taking too much or mishandling items, though these often had not materialised. Residents also frequently ask staff for assistance with selecting items. Staff noted that offering finger food helped residents who struggle with cutlery to eat more. They also highlighted the importance of portion control, as large portions could discourage residents from eating.

## Discussion

This review has shown how safe wandering in care homes can be facilitated. We developed a comprehensive theory encompassing context, strategies, mechanisms, and outcomes across five main programme theories: personalised care, monitoring, navigation, managing access, and hydration and nutrition. These elements illustrate the strategies that care homes can implement to help older adults to wander safely.

The literature emphasises the balance between safety and independence for individuals with dementia, a key aspect of our programme theory. Risk perception of wandering differs among institutions and individuals and is often influenced by adverse outcomes such as elopement and falls (7). While some studies focus on the dangers of wandering, it is equally important to highlight the positive aspects (12, 104). Neubauer & Liu (105) introduced the “Goldilocks Principle on Dementia and Wayfinding” to balance risk perception with the independence-safety spectrum. The framework emphasises enabling people with dementia to maintain autonomy while ensuring safety. It was developed to address concerns about elopement and getting lost in community settings, but can be applied to falls and injuries, which are a more significant concern for care home staff (25).

Many strategies employed across the programme theories raise potential ethical issues. Several strategies identified to enable wandering could also be employed to prevent wandering, including locking doors, using monitoring technology, and employing camouflage. These strategies are typically implemented to mitigate risks associated with residents leaving care homes or accessing unsafe areas. This review contextualises these strategies within the framework of preventing access to unsafe spaces while allowing residents the freedom to wander in secure and appropriate environments, highlighting the need to clearly define wandering support strategies and communicate them effectively across staff teams.

The use of wearable technology raises concerns about privacy and loss of autonomy and could result in discomfort and stigma for residents. The process of using such technologies should include a discussion between the resident, their family and the care home team, and informed consent should be received before implementation (15, 106).

Ongoing debate surrounds the ethics of exploiting perceptual deficits in people with dementia to keep them away from specific spaces, especially when visual hallucinations cause confusion or distress. While “compassionate deception” may support safe wandering (107), less intrusive options like limiting door visibility should be considered first. Strategies that deceive or restrict autonomy and freedom should be considered on a case-by-case basis, balancing risk with independence and prioritising alternatives like staff training; in line with the principles of least restrictive practice outlined in the Mental Capacity Act (2005) (108).

### Strengths and limitations

Strengths of the review include a broad and iterative search strategy, coverage of diverse and international evidence, detailed programme theories, and input from a lay advisory group. Findings were sense checked with key stakeholders. Studies were mainly conducted in Western countries, with one study from Asia. The inclusion of a paper written by some authors as a key text to inform searches represents a limitation. There was limited representation of the voices of people with dementia, with most evidence coming from care home staff. While capturing the views of those with dementia presents ethical and practical challenges, it is essential for gaining genuine insight (109, 110). Most literature focused on individual-level strategies, with little attention to how these work collectively in care homes.

### Implications for practice and research

Multiple approaches are available to enable residents to wander safely. Many small-scale strategies are easily implementable in care homes, whilst others require a shift in care culture or should be considered at the design stage. A proactive approach involving routine implementation of individualised non-pharmacological interventions could help reduce risks (111).

Residents describe how wandering provides a sense of purpose and enjoyment (12). A person-centred care approach can be applied to find the strategies that best suit each resident and their needs. Staff should get to know each resident well and anticipate their wandering behaviours and needs, including supporting their hydration and nutrition. A care home that encourages positive risk-taking in relation to wandering will provide residents with the chance to wander safely, with staff support when necessary (25). Assessment and regular review are required to ensure that strategies are appropriate for each individual (7), recognising that preferences may change as dementia progresses (74). Staff can walk with residents to offer reassurance and build relationships, however this is dependent on having sufficient staffing levels for one-to-one care.

Those building or redesigning a care home can take steps to help residents navigate independently. Design features include creating clear, distinct areas, using consistent design with varied colour schemes for orientation, and applying colour contrast to key features like handrails. Visual cues, landmarks, sensory stimuli, and accessible signage (with large print and pictograms) enhance navigation. Simple layouts with good visibility and monitoring technology foster autonomy while ensuring safety. Disguising areas to prevent unsafe entry may be effective, but raises ethical concerns (64).

Managers and staff must balance residents’ freedom to access spaces around the care home, while also ensuring their safety. Staff must possess the knowledge, skills, and support needed to balance resident preferences and safety in care homes (111). Training could focus on identifying why someone may be wandering and supporting residents in wandering safely. Managers and senior staff should support care staff to implement training in practice through shadowing and mentoring.

Future research should address identified evidence gaps, including examining the role of relatives in supporting safe wandering, and considering how strategies work when there are multiple residents who wander with different needs. Longitudinal research, incorporating the views of residents, relatives and staff should be conducted to understand how preferences and safety are balanced across care homes. Future research could explore how staffing and shift patterns affect strategies, and what the impact of supporting wandering is on staff. Evidence on how staff physically support residents and provide food and drink while they wander, is lacking and should be considered for future research.

## Conclusion

Allowing residents with dementia the freedom and autonomy to wander can prevent residents from feeling trapped and increase well-being. Physical and psychosocial strategies that create a facilitative environment, provide physical support, and creatively meet residents’ hydration and nutrition needs have been shown to support wandering in care homes. These strategies allow staff and managers to offer residents freedom while ensuring their safety, which leads to improved wellbeing for both residents and staff. Strategies to facilitate safe wandering should address the five programme theories.

## Data Availability

The data related to this review are available through the University of Sheffield ORDA (Figshare) repository at the following link:
https://doi.org/10.15131/shef.data.29339444.v1

## Acknowledgements

We wish to thank Ellie Little, Eleora Mansi, and Adrianna Montgomery, who supported with data extraction.

## Footnotes

1 Significant debate surrounds the language used to describe wandering, as the term implies restless movement without purpose. Alternatives such as ‘walking with purpose’ or ‘exploring the environment’ better reflect the intentions of individuals and help reduce stigma (Graham, 2017). Such a shift from a medical to a person-centred approach values individual needs, abilities, and identity (Kitwood, 1997). However, for consistency with current literature and terminology used by care staff, this review will use the term wandering.

