## Supplement 1 for "Strategies to support safe wandering in care homes for older adults – what works, for whom, and in which circumstances?: A realist synthesis"

Supplement 1: Search strategies

### (1) Initial search strategy

#### PubMed (all fields)

(("wander*"[All Fields] OR ("wandering behavior"[MeSH Terms] OR ("wandering"[All Fields] AND "behavior"[All Fields]) OR "wandering behavior"[All Fields]) OR "wayfind*"[All Fields] OR "walk*"[All Fields] OR ("walked"[All Fields] OR "walking"[MeSH Terms] OR "walking"[All Fields] OR "walks"[All Fields]) OR "orient*"[All Fields] OR "disorient*"[All Fields] OR ("orient"[All Fields] OR "orientability"[All Fields] OR "orientable"[All Fields] OR "orientate"[All Fields] OR "orientated"[All Fields] OR "orientates"[All Fields] OR "orientating"[All Fields] OR "orientation"[MeSH Terms] OR "orientation"[All Fields] OR "orientations"[All Fields] OR "orientation s"[All Fields] OR "orientation, spatial"[MeSH Terms] OR ("orientation"[All Fields] AND "spatial"[All Fields]) OR "spatial orientation"[All Fields] OR "oriented"[All Fields] OR "orientational"[All Fields] OR "orienting"[All Fields] OR "orients"[All Fields]) OR ("paced"[All Fields] OR "paces"[All Fields] OR "pacing"[All Fields] OR "pacings"[All Fields]) OR ("pacing clin electrophysiol"[Journal] OR "pace"[All Fields])) AND (("Care"[All Fields] AND "home*"[All Fields]) OR (("nursing"[MeSH Terms] OR "nursing"[All Fields] OR "nursings"[All Fields] OR "nursing"[MeSH Subheading] OR "nursing s"[All Fields]) AND "home*"[All Fields]) OR ("long term care"[MeSH Terms] OR ("long term"[All Fields] AND "Care"[All Fields]) OR "long term care"[All Fields] OR ("long"[All Fields] AND "term"[All Fields] AND "Care"[All Fields]) OR "long term care"[All Fields]) OR ("social support"[MeSH Terms] OR ("social"[All Fields] AND "support"[All Fields]) OR "social support"[All Fields] OR ("social"[All Fields] AND "Care"[All Fields]) OR "social care"[All Fields]) OR (("residential"[All Fields] OR "residentially"[All Fields]) AND "Care"[All Fields]) OR ("Care"[All Fields] AND ("facilities"[All Fields] OR "facility"[All Fields] OR "facility s"[All Fields])) OR ("residential facilities"[MeSH Terms] OR ("residential"[All Fields] AND "facilities"[All Fields]) OR "residential facilities"[All Fields]) OR ("nursing homes"[MeSH Terms] OR ("nursing"[All Fields] AND "homes"[All Fields]) OR "nursing homes"[All Fields]) OR ("long term care"[MeSH Terms] OR ("long term"[All Fields] AND "Care"[All Fields]) OR "long term care"[All Fields] OR ("long"[All Fields] AND "term"[All Fields] AND "Care"[All Fields]) OR "long term care"[All Fields]) OR ("residential facilities"[MeSH Terms] OR ("residential"[All Fields] AND "facilities"[All Fields]) OR "residential facilities"[All Fields]) OR ("homes for the aged"[MeSH Terms] OR ("homes"[All Fields] AND "aged"[All Fields]) OR "homes for the aged"[All Fields]) OR ("assisted living facilities"[MeSH Terms] OR ("assisted"[All Fields] AND "living"[All Fields] AND "facilities"[All Fields]) OR "assisted living facilities"[All Fields])) AND ("alzheimer disease"[MeSH Terms] OR ("alzheimer"[All Fields] AND "disease"[All Fields]) OR "alzheimer disease"[All Fields] OR ("dementia"[MeSH Terms] OR "dementia"[All Fields] OR "dementias"[All Fields] OR "dementia s"[All Fields]) OR "alzheimer*"[All Fields])) AND (english[Filter])

Translations

Wandering Behavior: "wandering behavior"[MeSH Terms] OR ("wandering"[All Fields] AND "behavior"[All Fields]) OR "wandering behavior"[All Fields]

Walking: "walked"[All Fields] OR "walking"[MeSH Terms] OR "walking"[All Fields] OR "walks"[All Fields]

Orientation: "orient"[All Fields] OR "orientability"[All Fields] OR "orientable"[All Fields] OR "orientate"[All Fields] OR "orientated"[All Fields] OR "orientates"[All Fields] OR "orientating"[All Fields] OR "orientation"[MeSH Terms] OR "orientation"[All Fields] OR "orientations"[All Fields] OR "orientation's"[All Fields] OR "orientation, spatial"[MeSH Terms] OR ("orientation"[All Fields] AND "spatial"[All Fields]) OR "spatial orientation"[All Fields] OR "oriented"[All Fields] OR "orientational"[All Fields] OR "orienting"[All Fields] OR "orients"[All Fields]

pacing: "paced"[All Fields] OR "paces"[All Fields] OR "pacing"[All Fields] OR "pacings"[All Fields]

pace: "Pacing Clin Electrophysiol"[Journal:__jid7803944] OR "pace"[All Fields]

nursing: "nursing"[MeSH Terms] OR "nursing"[All Fields] OR "nursings"[All Fields] OR "nursing"[Subheading] OR "nursing's"[All Fields]

long-term care: "long-term care"[MeSH Terms] OR ("long-term"[All Fields] AND "care"[All Fields]) OR "long-term care"[All Fields] OR ("long"[All Fields] AND "term"[All Fields] AND "care"[All Fields]) OR "long term care"[All Fields]

social care: "social support"[MeSH Terms] OR ("social"[All Fields] AND "support"[All Fields]) OR "social support"[All Fields] OR ("social"[All Fields] AND "care"[All Fields]) OR "social care"[All Fields]

residential: "residential"[All Fields] OR "residentially"[All Fields]

facilities: "facilities"[All Fields] OR "facility"[All Fields] OR "facility's"[All Fields]

residential facilities: "residential facilities"[MeSH Terms] OR ("residential"[All Fields] AND "facilities"[All Fields]) OR "residential facilities"[All Fields]

Nursing Homes: "nursing homes"[MeSH Terms] OR ("nursing"[All Fields] AND "homes"[All Fields]) OR "nursing homes"[All Fields]

Long-Term Care: "long-term care"[MeSH Terms] OR ("long-term"[All Fields] AND "care"[All Fields]) OR "long-term care"[All Fields] OR ("long"[All Fields] AND "term"[All Fields] AND "care"[All Fields]) OR "long term care"[All Fields]

Residential Facilities: "residential facilities"[MeSH Terms] OR ("residential"[All Fields] AND "facilities"[All Fields]) OR "residential facilities"[All Fields]

Homes for the Aged: "homes for the aged"[MeSH Terms] OR ("homes"[All Fields] AND "aged"[All Fields]) OR "homes for the aged"[All Fields]

Assisted Living Facilities: "assisted living facilities"[MeSH Terms] OR ("assisted"[All Fields] AND "living"[All Fields] AND "facilities"[All Fields]) OR "assisted living facilities"[All Fields]

Alzheimer Disease: "alzheimer disease"[MeSH Terms] OR ("alzheimer"[All Fields] AND "disease"[All Fields]) OR "alzheimer disease"[All Fields]

dementia: "dementia"[MeSH Terms] OR "dementia"[All Fields] OR "dementias"[All Fields] OR "dementia's"[All Fields]

#### Cochrane Library (title abstract keyword)

Wander* OR Wandering Behavior OR wayfind* OR walk* OR Walking OR orient* OR disorient* OR Orientation OR pacing OR pace in Title Abstract Keyword AND Care home* OR nursing home* OR long-term care OR social care OR residential care OR care facilities OR residential facilities OR Nursing Homes OR Long-Term Care OR Residential Facilities OR Homes for the Aged OR Assisted Living Facilities in Title Abstract Keyword AND Alzheimer Disease OR dementia OR Alzheimer* in Title Abstract Keyword - with Cochrane Library publication date Between Jan 2005 and Dec 2024, in Cochrane Reviews, Trials, Editorials (Word variations have been searched)

#### PsycINFO (multipart (.mp))

1	(Wander* or Wandering Behavior or wayfind* or walk* or Walking or orient* or disorient* or Orientation or pacing or pace).mp.

2	(Care home* or nursing home* or long-term care or social care or residential care or care facilities or residential facilities or Nursing Homes or Long-Term Care or Residential Facilities or Homes for the Aged or Assisted Living Facilities).mp.

3	(Alzheimer Disease or dementia or Alzheimer*).mp.

4	1 and 2 and 3

5	limit 4 to (english language and yr="2005 -Current")

#### CINAHL (all fields)

( Wander* OR Wandering Behavior OR wayfind* OR walk* OR Walking OR orient* OR disorient* OR Orientation OR pacing OR pace ) AND ( Care home* OR nursing home* OR long-term care OR social care OR residential care OR care facilities OR residential facilities OR Nursing Homes OR Long-Term Care OR Residential Facilities OR Homes for the Aged OR Assisted Living Facilities ) AND ( Alzheimer Disease OR dementia OR Alzheimer* )

Limiters - Publication Date: 20050101-20241231

Expanders - Apply equivalent subjects

Narrow by Language: - English

Search modes - Proximity

ProQuest Dissertations & Theses Global - anywhere except full text (NOFT)

noft(Wander* OR Wandering Behavior OR wayfind* OR walk* OR Walking OR orient* OR disorient* OR Orientation OR pacing OR pace) AND noft(Care home* OR nursing home* OR long-term care OR social care OR residential care OR care facilities OR residential facilities OR Nursing Homes OR Long-Term Care OR Residential Facilities OR Homes for the Aged OR Assisted Living Facilities) AND noft(Alzheimer Disease OR dementia OR Alzheimer*) AND la.exact("English") AND pd(>20041231)

#### Scopus (title, abstract, keywords)

( TITLE-ABS-KEY ( wander* OR wandering AND behavior OR wayfind* OR walk* OR walking OR orient* OR disorient* OR orientation OR pacing OR pace ) AND TITLE-ABS-KEY ( care AND home* OR nursing AND home* OR long-term AND care OR social AND care OR residential AND care OR care AND facilities OR residential AND facilities OR nursing AND homes OR long-term AND care OR residential AND facilities OR homes AND for AND the AND aged OR assisted AND living AND facilities ) AND TITLE-ABS-KEY ( alzheimer AND disease OR dementia OR alzheimer* ) ) AND PUBYEAR > 2004

#### NIHR Journals Library

Wandering; wander

### (2) Targeted searched focused on prioritised IPTs

#### IPT10 - Residents are adequately nourished and/or hydrated

##### Scite

nutrition and wandering and dementia

##### Litsense

nutrition and wandering and dementia

##### PubMed

15.10.24 search:

- ((Wander* OR Wandering Behavior OR wayfind* OR walk* OR Walking OR orient* OR disorient* OR Orientation OR pacing OR pace) AND (Alzheimer Disease OR dementia OR Alzheimer*) AND (nutrition or nourish* or hydrat*)) - 354 hits - no relevant additional records
- nourishment and wandering and dementia - 1 hit
- hydration and wandering and dementia - 0 hits
- (food or drink) and wandering and dementia - 14 hits
- ((Wander* OR Wandering Behavior OR wayfind* OR walk* OR Walking OR orient* OR disorient* OR Orientation OR pacing OR pace) AND (Alzheimer Disease OR dementia OR Alzheimer*) AND (nutrition or nourish* or hydrat* or food or drink)), limited to title/abstract - 128 hits

##### Litsense

15.10.24 search:

- nourishment and wandering and dementia
- hydration and wandering and dementia
- food and wandering and dementia
- drink and wandering and dementia

28.10.24 search:

- Energy and wandering and dementia
- Fatigue and wandering and dementia

29.10.24 search:

- Care home residents with dementia are adequately nourished and/or hydrated when wandering

##### Scite

24.11.24 search:

- nourishment and wandering and dementia (boolean mode)
- hydration and wandering and dementia (boolean mode)
- food and wandering and dementia (boolean mode)
- drink and wandering and dementia (boolean mode)
- Energy and wandering and dementia (boolean mode)
- Fatigue and wandering and dementia (boolean mode)

#### IPT4 - Staff understanding of why each resident wanders

Combination of staff understanding, wandering and dementia

##### Scite

Staff understanding and wandering and dementia

##### Litsense

28.10.24 search:

- Staff and understand and wandering and dementia
- Staff and knowledge and wandering and dementia
- Staff and education and wandering and dementia
- Staff and training and wandering and dementia
- Life history and wandering and dementia
- Staff and reason and wandering and dementia

##### Pubmed

28.10.24 search:

- ((Wander* OR Wandering Behavior OR wayfind* OR walk* OR Walking OR orient* OR disorient* OR Orientation OR pacing OR pace) AND (Alzheimer Disease OR dementia OR Alzheimer*) AND ((staff OR carer) AND understand*)) - 202 hits - from eyeballing titles of the first 30, they don’t look particularly relevant.

##### Litsense

29.10.24 searches:

- Care home staff understand why each resident wanders- nothing new/relevant

##### Scite

24.11.24 search:

- Staff understanding and wandering and dementia (boolean mode)

27.11.24 search:

- Care home staff understand why each resident wanders

28.11.24 search:

- Staff and understand and wandering and dementia
- Staff and knowledge and wandering and dementia
- Staff and education and wandering and dementia
- (29.11.24) Life history and wandering and dementia
- Staff and reason and wandering and dementia

#### IPT7 - Residents can’t access dangerous spaces

##### Scite

- Camouflage and dementia and wandering

##### Pubmed

28.10.24 search:

- ((Wander* OR Wandering Behavior OR wayfind* OR walk* OR Walking OR orient* OR disorient* OR Orientation OR pacing OR pace) AND (Alzheimer Disease OR dementia OR Alzheimer*) AND (camouflag*))

##### Pubmed

8.11.24 search:

- ((Wander* OR Wandering Behavior OR wayfind* OR walk* OR Walking OR orient* OR disorient* OR Orientation OR pacing OR pace) AND (Alzheimer Disease OR dementia OR Alzheimer*) AND (disguis*))

##### Litsense

8.11.24 search:

- disguised and wandering and dementia
- Camouflage and dementia and wandering

##### Scite

29.11.24 search:

- Camouflage and dementia and wandering
- Disguised and wandering and dementia

#### IPT3 - Staff detect when residents need support

##### Litsense

8.11.24 search:

- Support and wandering and dementia
- Staff detect when residents need support with wandering in care homes

##### Scite

29.11.24 search:

- Support and wandering and dementia
- Staff detect when residents need support with wandering in care homes

1.12.24 search:

- Noticing and wandering and dementia
- Awareness and wandering and dementia

#### IPT9 - Residents are supported physically

##### Scite

2.12.24 search:

- Supporting and wandering and dementia (Boolean mode)
- “Walking with” and wandering and dementia (Boolean mode)
- “Walking aid” and wandering and dementia (Boolean mode)
- Accompanying AND wandering AND dementia

##### PubMed

3.12.24 search:

- (Wander* OR Wandering Behavior OR wayfind* OR walk* OR Walking OR orient* OR disorient* OR Orientation OR pacing OR pace) AND (Alzheimer Disease OR dementia OR Alzheimer*) AND (support* OR "walking aid" OR "walking with") - 1354 hits

##### Litsense

10.11.24 search:

- Person-centred care and wandering and dementia
- Person-centered care and wandering and dementia

##### Scite

6.12.24 search:

- Person-centred care and wandering and dementia
- Person-centered care and wandering and dementia

#### Searches around a key paper (Griffiths et al., 2024)

Google Scholar (8.11.24): no citations, 100 ‘related articles’

Scite (20.11.24): related articles.

### (3) Grey literature searches

#### Documenting the search and records found

| **Name of resource (and website)** | **Date searched** | **Date of last access (if different)** | **Pathway followed, e.g. browsed headings/searched site/database within website (use separate lines for the different types of searches)** | **Notes and links to relevant resources** |
| --- | --- | --- | --- | --- |
| British Geriatrics Society <https://www.bgs.org.uk/> | 18.11.24 |  | Resources → Search our clinical and research resources  Searched “dementia and wandering”  2 documents found  Searched “wandering” - same two documents found |  |
| British Society for Gerontology <https://www.britishgerontology.org/> | 18.11.24 |  | Publications → BSG Reports and Submissions  Only one report on there, doesn’t look relevant  Publications → Generations Review - The Newsletter  Looked through:  June 2017  October 2017 | There are links to these journals - would they have been indexed to PubMed?  Ageing & Society - The Journal  Journal of Global Ageing  Journal of Population Ageing  Canadian Journal on Ageing  International Journal of Care and Caring |
| Care Quality Commission <https://www.cqc.org.uk/> | 22.11.24 |  | Publications →  Searched “dementia and wandering” - 40 pages of results  Searched long documents for ‘wander’ and ‘walk’ using find function  Searched “dementia wandering” - 6 pages of results  Searched “wander” - only one record, which came up under previous searches | Seems to be including the word ‘and’ in the search. |
| Care England [https://www.careengland.org.uk](https://www.careengland.org.uk/) | 22.11.24 |  | Resources and guidance → Resources and guidance  Searched “dementia wander” - 4 pages of results  News and press  Searched “dementia wander” - 13 pages of results | Nothing of relevance    Nothing of relevance |
| The Care Forum <https://www.thecareforum.org/> | 22.11.24 |  | Research and Projects [no search function just a list of research resources in thumbnails over two pages]  Policy Voice [no search function just a list of policy resources in thumbnails over one page]    News → Latest News → Blog [no search function, 8 pages of thumbnails]  News → Latest News → Care Sector News [no search function, 27 pages of thumbnails]  News → Latest News → Reports and resources [no search function, one page of thumbnails]  News → NCF Publications [no search function, one page of thumbnails]  Website search function: “dementia and wandering” - one resource, title suggests not relevant.  “Dementia and wander” - two resources, titles suggest not relevant.  “Dementia wander” - same two results. | Nothing of relevance    There might be something within one of the documents on this Responses to Consultations and Calls for Evidence page, although it seems unlikely: <https://www.nationalcareforum.org.uk/voice/responses-to-consultations-and-calls-for-evidence/>  Looked through - nothing of relevance  Nothing of relevance    Nothing of relevance    Nothing of relevance    Nothing of relevance    Nothing of relevance    Nothing of relevance    Nothing of relevance |
| National Care Association <https://nationalcareassociation.org.uk/> | 22.11.24 |  | Resources → Publications  One listed - not relevant  Resources → Useful links  2-page list - nothing relevant  News & Events → Latest news  2-page list - nothing relevant | NOTE: there is a member area that I can’t access |
| The Care Forum <https://www.thecareforum.org/> | 22.11.24 |  | Our Projects (On home page, scrolled down) → Reports & Influence [nothing relevant]  News → Latest news  [178 pages - checked first 4 and nothing relevant, very general]  No way of searching whole site but seems very broad in focus |  |
| Care choices <https://www.carechoices.co.uk/> | 22.11.24 |  | Publications  Not relevant (relate to choosing care - guides)  Blog  [28 pages of lists/thumbnails of blog posts] | Blogs - this blog post mentions providing finger food while on the move and has a link - not much detail at all but may be useful: <https://www.carechoices.co.uk/blog/dementia-and-nutrition/>  This blog post has some advice on supporting walking in people with dementia - does not mention setting but does not appear to be setting-dependent: <https://www.carechoices.co.uk/blog/supporting-a-person-with-dementia-to-walk/>  This might be relevant - on personalising zimmer frames <https://www.carechoices.co.uk/blog/zinged-zimmers-reduce-falls/>  This blog post has something on orientation (although not very much and only tangentially linked to wandering): <https://www.carechoices.co.uk/blog/street-signs-help-dementia-residents-navigate-their-care-home/>  Small mention of going out for a walk in context of high-quality person-centred care <https://www.carechoices.co.uk/blog/outstanding-staff-praised-cqc-report/> |
| Department of Health and Social Care <https://www.gov.uk/government/organisations/department-of-health-and-social-care> | 22.11.24 |  | Research and statistics → See all research and statistics - 718 results  Searched “dementia” - 25 results - nothing of relevance  Searched “wandering” and “wander” - only one hit, duplicated from previous.  News and communications → See all news and communications  Searched “dementia” - 324 results  Searched “dementia and wandering” - 326 results. Skimmed titles, nothing obviously relevant.  Policy papers and consultations → See all Policy papers and consultations  Searched “dementia” - 71 results - nothing relevant |  |
| Social Care Institute for Excellence <https://www.scie.org.uk/> | 3.12.24 |  | Care themes → Personalised and person-centred care → Meeting the needs of every individual → Find out more  About SCIE → Annual report → 2022/23  About SCIE → Featured articles and opinions  About SCIE → News  About SCIE → Events  Search box: dementia; wander; wandering | Nothing of relevance |
| Skills for care <https://www.skillsforcare.org.uk/> | 3.12.24 |  | News and events → Events and networks  News and events → News  News and events → Blogs and articles  Search box: dementia; wander; wandering | Nothing of relevance |
| Care Inspectorate Wales <https://www.careinspectorate.wales/> | 3.12.24 |  | Our reports → Chief Inspector’s Annual Report → 2023-24, 2022-23, 2021-22, 2020-21, 2019-20, 2018-19  News  Search box: dementia; wander; wandering | Nothing of relevance |
| Social care Wales <https://socialcare.wales/> | 4.12.24 |  | Resources and guidance → Improving care and support → people with dementia  Research, data and innovation → Workforce reports  Research, data and innovation → Linked data research  Research, data and innovation → Curated research  Search box: dementia; wander; wandering | Nothing of relevance |
| Alzhiemers Society <https://www.alzheimers.org.uk/> | 4.12.24 |  | Research → Dementia research news (focuses on diagnosis and treatments)  Search box: wander; wandering | Nothing of relevance |
| Alzhiemers Research UK <https://www.alzheimersresearchuk.org/> | 4.12.24 |  | Research → About our research → Our projects and initiatives (categorised as ‘treat’, ‘diagnose’ and ‘prevent’, none of which fit, so did not explore the categories further)  Search box: wander; wandering | Nothing of relevance |
| Age UK <https://www.ageuk.org.uk/> |  |  | Our impact → Policy and research → Reports and publications → Reports and briefings → Care and support  Our impact → Policy and research → Reports and publications → Reports and briefings → Health and wellbeing  Our impact → Policy and research → Reports and publications → Consultation responses → Care and support  Our impact → Policy and research → Reports and publications → Consultation responses → Health and wellbeing  Our impact → Policy and research → Reports and publications → Evaluation reports → Dementia services  Search box: dementia; wander; wandering |  |
| Health Building Note 08-02 <https://www.england.nhs.uk/publication/dementia-friendly-health-and-social-care-environments-hbn-08-02/> | 4.12.24 |  | Single document - read through. | The document contains lots of guidance relating to building design for orientation, although it might be challenging to extract dyads/triads - they would be assumed. |
| Dementia Voices <https://www.dementiavoices.org.uk/> | 5.12.24 |  | DEEP resources → Publications by DEEP groups  DEEP resources → DEEP gatherings and events → Gatherings reports  DEEP news → Our DEEP news  Search box: wander; wandering | Nothing of relevance |
| NHS website <https://www.nhs.uk/> | 6.12.24 |  | Search box: dementia; wander; wandering | Nothing of relevance |
| SIGN <https://www.sign.ac.uk/our-guidelines/> | 6.12.24 |  | List of guidelines (looked through)  Search box: dementia; wander; wandering | Nothing of relevance |
| Local authorities (Google) | 6.12.24 |  | Google search:  "local authority" and dementia and wandering  "local authority" and dementia and wander  council and dementia and wander  Local Government Association website (<https://www.local.gov.uk/>)  Search box: dementia and wandering; dementia and wander | There is a tiny bit in this news article about detecting when people are getting up in the night but unclear if it is to support wandering <https://www.theguardian.com/society/2022/dec/30/dementia-village-in-warwick-is-a-pioneer-in-person-centred-care> |
| Dementia Pathfinders <https://www.dementiapathfinders.org/> | 6.12.24 |  | News → Blog  Search box: wander; wandering | Nothing of relevance |
| NICE <https://www.nice.org.uk/> | 6.12.24 |  | Search box: dementia and wandering; dementia and wander; dementia |  |

#### Documenting the initial screening process

| **Name of resource** | **No. of promising documents** | **Number scanned** | **Approach to screening, e.g. title, then abstract/full text OR first 100 ranked by relevance** | **Additional notes** |
| --- | --- | --- | --- | --- |
| British Geriatrics Society <https://www.bgs.org.uk/> | 0 | 1 | Only two resources identified. One did not look relevant by title.  Examined full text of “End of Life Care in Frailty: Dementia” guidance - nothing relevant in there. | Nothing of relevance |
| British Society for Gerontology <https://www.britishgerontology.org/> | 0 | 0 | Nothing relevant was returned |  |
| Care Quality Commission <https://www.cqc.org.uk/> | 0 | 18 | Looked at titles and extracts of relevant resources. Any that looked like it might have something on wandering in dementia were examined in full. Large documents were searched for “wander” and “walk” using the find function. | Nothing of relevance |
| Care England [https://www.careengland.org.uk](https://www.careengland.org.uk/) | 0 | 22 | Looked at titles and extracts of relevant resources. Any that looked like it might have something on wandering in dementia were examined in full. Large documents were searched for “wander” and “walk” using the find function. | Nothing of relevance |
| The Care Forum <https://www.thecareforum.org/> | 0 | 61 | Looked at titles and extracts of relevant resources. Any that looked like it might have something on wandering in dementia were examined in full. Large documents were searched for “wander” and “walk” using the find function. | Nothing of relevance |
| National Care Association <https://nationalcareassociation.org.uk/> | 0 | 1 | Looked at titles and extracts of relevant resources. Any that looked like it might have something on wandering in dementia were examined in full. | Nothing of relevance |
| The Care Forum <https://www.thecareforum.org/> | 0 | 0 | Nothing screened | Nothing of relevance |
| Care choices <https://www.carechoices.co.uk/> | 5 | 28 | Looked at titles and extracts of relevant resources. Any that looked like it might have something on wandering in dementia were examined in full. | Five blog posts with a bit of relevant information in. See links in table above. |
| Department of Health and Social Care <https://www.gov.uk/government/organisations/department-of-health-and-social-care> | 0 | 10 |  |  |
| Social Care Institute for Excellence <https://www.scie.org.uk/> | 0 | 12 | Annual report: looked through the 2022/23 report (nothing of relevance)  Featured articles and opinions: skimmed titles, read any articles that looked potentially relevant  News: skimmed titles, read any articles that looked potentially relevant  Events: skimmed titles (none relevant)  Search box: skimmed titles for each search, read any articles that looked potentially relevant | Nothing of relevance |
| Skills for care <https://www.skillsforcare.org.uk/> | 0 | 4 | Skimmed titles for each search, read any articles that looked potentially relevant.  Blogs and articles - screened the first 25 pages. |  |
| Care Inspectorate Wales <https://www.careinspectorate.wales/> | 0 | 7 | Reports - skimmed, went to relevant section (care homes) and searched document using ‘find’ function for “dementia”, “wander” and “walk”.  News - skimmed titles in list (first 10 pages - back to May 2023).  Search box: skimmed titles for each search, read any articles that looked potentially relevant. | Nothing of relevance |
| Social care Wales <https://socialcare.wales/> | 0 | 15 | Skimmed titles of documents and pages and read any in full that looked relevant. | Nothing of relevance |
| Alzhiemers Society <https://www.alzheimers.org.uk/> | 0 | 2 | Skimmed titles of documents and pages and read any in full that looked relevant. | Nothing of relevance |
| Alzhiemers Research UK <https://www.alzheimersresearchuk.org/> | 0 | 0 | Skimmed titles of documents and pages and read any in full that looked relevant. | This all seems to be clinical. None of the news stories seem to be relevant. |
| Age UK <https://www.ageuk.org.uk/> | 0 | 3 | Skimmed titles of documents and pages and read any in full that looked relevant. | Nothing of relevance |
| Health Building Note 08-02 (Dementia Friendly Health Building Note) <https://www.england.nhs.uk/publication/dementia-friendly-health-and-social-care-environments-hbn-08-02/> | 1 | 1 | Single document - read through. | The document contains lots of guidance relating to building design for orientation, although it might be challenging to extract dyads/triads - they would be assumed |
| Dementia Voices <https://www.dementiavoices.org.uk/> | 0 | 46 | Skimmed titles of documents and pages and read any in full that looked relevant. Searched long documents using ‘find’ function for “wander” and “walk”.  Newsletters screened back to January 2021 - seem focused on community living and nothing of relevance. | Nothing of relevance |
| NHS website <https://www.nhs.uk/> | 0 | 0 | Screened titles, nothing of relevance | Nothing of relevance |
| SIGN <https://www.sign.ac.uk/our-guidelines/> | 0 | 1 | Screened titles of guidelines, read through full texts of any that looked relevant | Nothing of relevance |
| Local authorities (Google) | 0 | 13 | Screened titles on first four pages, looked at full text/webpage of anything relevant | Nothing of relevance |
| Dementia Pathfinders <https://www.dementiapathfinders.org/> | 0 | 1 | Screened titles of blog posts, read through any posts that looked relevant | Nothing of relevance |
| NICE <https://www.nice.org.uk/> | 0 | 5 | Screened titles of guidelines and quality standards in search results and looked at full texts of any that looked relevant. Searched long documents using ‘find’ function for “wander”, “walk”, “mov”, “amb”, “pace” and “pacing”. | Nothing of relevance |

#### List of organisations to search

**Organisation**

British Geriatrics Society <https://www.bgs.org.uk/>

British Society for Gerontology <https://www.britishgerontology.org/>

Care Quality Commission <https://www.cqc.org.uk/>

Care England <https://www.careengland.org.uk/>

National Care Forum <https://www.nationalcareforum.org.uk/>

National Care Association <https://nationalcareassociation.org.uk/>

The Care Forum <https://www.thecareforum.org/>

Care choices <https://www.carechoices.co.uk/>

Department of Health and Social Care <https://www.gov.uk/government/organisations/department-of-health-and-social-care>

Social Care Institute for Excellence <https://www.scie.org.uk/>

Skills for care <https://www.skillsforcare.org.uk/>

Care Inspectorate Wales <https://www.careinspectorate.wales/>

Social care Wales <https://socialcare.wales/>

Alzheimer's Society <https://www.alzheimers.org.uk/>

Alzheimer's Research UK <https://www.alzheimersresearchuk.org/>

Age UK <https://www.ageuk.org.uk/>

Health Building Note 08-02 (Dementia Friendly Health Building Note) <https://www.england.nhs.uk/publication/dementia-friendly-health-and-social-care-environments-hbn-08-02/>

Dementia Voices <https://www.dementiavoices.org.uk/>

NHS website <https://www.nhs.uk/>

SIGN <https://www.sign.ac.uk/our-guidelines/>

Local authorities - search Google for “local authorities” and “wander”, check first four pages

Dementia Pathfinders <https://www.dementiapathfinders.org/>

NICE <https://www.nice.org.uk/>
