## Supplement 2 for "Strategies to support safe wandering in care homes for older adults – what works, for whom, and in which circumstances?: A realist synthesis"

Supplement 2: Relevance, richness and rigour judgements

### Table S2.1: Judgements of relevance, richness and rigour for evidence sources included in the realist synthesis

| **First author, date** | **Relevance** | **Richness** | **Rigour** |
| --- | --- | --- | --- |
| **Evidence sources identified through initial database searches** | | | |
| Alam 2023 | Relevant. This paper talks about design elements that support wayfinding. Some data extracted relates to context and mechanisms but very little. There is little about outcome. This is more staff perspective rather than describing what the residents experience. | High. There is information on different design elements to support wayfinding, although this is the staff's perspective rather than the residents' experience. Focuses mainly on one IPT with a quick mention on two other but does not discuss the context or mechanism of those IPTS | Only focused on one area of the USA in two homes so lacks representative. We have no context on what these settings are like to know how this affects staff views. The staff were picked by the head nurse therefore the sample could have selection bias. |
| Anderson 2023 | High relevance - Information relating to various types of camouflage and possible to extract dyads | Moderate - Some detail relating to findings of relevant included studies | Details of search reported but not of other review methods. Quality assessment not undertaken. Unclear how studies were selected. |
| Anonymous 2007 | Somewhat relevant. The article provides recommendations to prevent wandering or elopement but this is not a study | Low. This is not a study it is more of a commentary piece. Only a few bits extracted only about the interventions, and a little on context nothing for mechanisms or outcomes | Low. This is a commentary piece. |
| Apple 2015 | Moderate- some information extracted but wandering was not the main topic | Moderate- some date extracted on context and outcomes but none on mechanisms | Moderate- only one care home studied although the researcher did attempt to work with other care homes but they declined, small sample group, strong methodology use three approaches for triangulation |
| Barrett 2020 | Very relevant, the paper touches on a number of IPTs; there is a little discussion about context and outcomes but very minimal, if any, on the mechanisms. Most of the outcomes are more about how certain strategies help staff (e.g. time, stress) rather than outcomes for the resident. | Moderate richness detail on multiple IPT but mainly discussion of strategies rather than being able to link strategies to context, mechanisms and outcomes. | Moderate The survey had high response rates from a number of care homes but all care homes was from the same organisation and the sample was self-selected. There was not a lot of information about the care homes. The data extracted is the perspective of care staff rather than the actual experience of residents. The case studies was a small sample. |
| Benbow 2017 | High relevance- a lot of data extracted for a number of IPTs, has a lot of context and some outcomes but no mechanisms | Moderate richness- was a commentary on other people’s work so richness is limited but a lot of data extracted, a lot for context, some for outcomes, none for mechanisms | Moderate - it was a commentary piece that discusses others work so rigour would be dependent of the work that they discuss |
| Brush 2008 | Moderate- Does discuss wayfinding but is discussing others work that we have already included. Information extracted is limited | Moderate- some data extracted but little context, mechanisms or outcomes. The paper discusses other work in limited description | Low- is a commentary pieces no information on the methods of the papers discussed |
| Brush 2015 | High relevance- explores IPT 6 wayfinding, also explores residents’ preferences to colour and pictures | Moderate- some information extracted on context and outcomes, little to none on mechanisms | Moderate- detailed and appropriate mechanisms, only two care homes with no information on the care home design and the recruitment strategy, no information on recruitment of residents |
| Caffo 2014 | High relevance- compares two interventions for PT5, some context, outcomes and mechanisms presented | Moderate- Some information for context outcomes and mechanisms | Moderate- Detailed methodology, uses inter-rater reliability, no information on recruitment |
| Cheung 2021 | Moderate- may be suitable for a counter theory for IPT 3. Looks more at preventing than supporting | Moderate- some context, no outcomes of mechanisms | Moderate- only one care home, method is unclear |
| Cohen-Mansfield 1998 | Moderate- some information for PT 8 some context and outcome, no mechanisms | Moderate- Some context and outcomes, no mechanisms | Moderate- small sample size, only one care home, short intervention time, did not account for confounding variables |
| Detweiler 2008 | High- discusses a wandering garden that addresses PT8 | High - Rich data on how the intervention works and in which contexts | Moderate- no information on the setting, recruitment and authors pick out some limitations to the study design |
| Detweiler 2009 | High- very relevant for IPT8 | high- rich data on IPT8 for context, mechanisms and outcomes | moderate. The small size of the sample, as well as its convenience nature, is a serious limitation. By using 2 years of observational data, the researchers attempted to have the participants serve as their own controls. A preferred experimental design would include a separate matched control |
| Dewing 2011 | Low- do discuss some IPT but very limited data extracted no outcomes, context or mechanisms | Low- very limited data extracted no outcomes, context or mechanisms. | Low- this is a commentary piece and do not discuss the methodology of any of the studies discussed. |
| Dickinson 1998 | Very relevant, the paper touches on a number of IPTs especially on accessing dangerous areas, some information could be used as a counter theory as could be considered preventing rather than supporting. Each participant is discussed individual allowing for data to be extracted who the strategies work for. There is a lot of data on the mechanisms and some on the context | Very rich, can extract context, mechanisms and outcomes. However there were only 8 participants and some of the participants date was not assess for varying reason so the information is from a small number. | Low. One institution only eight residents and several were not included in the analysis. no information on how the care home and residents were recruited. |
| Dickinson 1995 | Moderate relevance- discusses one IPT but the intervention is discussed in other papers. Could be used as a counter theory. | Moderate very short paper so little data extracted some data on mechanisms no information on outcomes or context | Low short paper so minimal information on methods no information on recruitment of the care home or participants. |
| Duffy 2019 | relevant- extracted data for a number of IPTs | moderate- this was a commentary pieces. A few bits extracted put mainly about the intervention, no context, mechanisms and outcomes. | Low. This is a commentary piece. |
| Faith 2014 | High relevance, talks about several IPTs in great detail. Discussed each care home and participant individually so able to determine context | High richness, a lot of data was extracted. There was data for outcomes, context and some information on mechanisms. | High rigour, explains the methods and recruitment in detail. the recruitment was rigorous to determine appropriate care homes although they were unable to observe all layout types. In one care home only one participant which reduces the rigour. |
| Fine 2015 | Low - includes design principles about orientation and architectural design | Low - no empirical data on mechanisms or outcomes | Low - MA thesis describing hypothetical care home design project, unclear what, methods and theory, support claims in the paper |
| Gibson 2004 | Moderate- some information on wayfinding but information very limited. This was a very short paper so little on context or mechanisms. | Moderate- this was a very short paper so only so much information could be extracted. Discusses wayfinding and intruding on others but didn't really focus on the IPTs in detail to explain it | Moderate- unable to generalise the study results as it is a small sample size. Only one care home that specialised in veterns. the study was conducted based on convience sample. |
| Griffiths 2024 | Highly relevant across several IPTs, possible to extract at least dyads | High. Very rich data with quotations | High. Good methodological quality, clear sampling, data collection and analysis methods and reflexivity |
| Gulwadi 2013 | Moderate- discusses an intervention relating to PT6 but only limited information on outcomes and context, no information mechanisms | Moderate- information limited. | Explain why these care home wards were selected but no information on how this care home is similar or different from other care homes. Detailed methods. |
| Heward 2022 | High relevance- talking about several IPT, was able to extract context on the care homes but no discussion of who stratergies work for, some outcomes mention but no mention of mechanisms | Moderate richness- detail on multiple IPT but more around context than outcomes or mechanisms. A lot of information about spatial orientation but may not fit fully into the IPTs | Moderate richness- detailed methods and recruitment. Recruitment of care homes was a purposive sampling of care homes that have previously taken part in research so may not represent all care homes. they excluded care homes part of organisations due to changes be of a result of the organisation not managers. small sample size |
| Hirst 1989 | Low- not really relevant some information for 10 and 4 mainly looks at preventing | Low- only a few lines extracted | Low- commentary piece |
| Holthe 2018 | Low- not really relevant only 3 bits of data extracted and only studies that looked at interventions for safe wandering | Low- no data extracted on context mechanisms or outcomes. | Moderate- assessed the quality of the studies but chose not to exclude the low rating studies, there could be some bias on their search strategies |
| Hussain 1987 | High relevance - Information relating to various configurations of floor markings and possible to extract dyads | Low to moderate - quantitative data but some information on mechanism and context | Moderate. Experiment undertaken scientifically - the only variable manipulated was the presence/type of marking and behaviour was observed by at least one observer with a framework/criteria. "Fifty of the 139 total opportunities were observed by both observers. Interobserver agreement as to crossing and not crossing was 100%." However, only undertaken in one location, which was more clinical in nature, and a long time ago (1980s). Observers were not blinded, however. Only 8 PWD took part and not all approached the door the same number of times. |
| Ilem 2018 | Relevant, talks about wayfinding in great detail. Each participant is described individually so can extract data on who the intervention works for. Data extracted on context and mechanisms | Moderate richeness only discusses one IPT each individual discusses seperate with some data extracted on outcomes and mechanisms | Moderate rigour no discussion on how the care homes were recruited and only two care homes took place. THey outline some issues with the methods used including that language impairments may have impacted results. Only one male. THey did not account for confounding variables . |
| Innes 2011 | Moderate relevance discusses several IPT but only a few bits of data was extracted. | Moderate richness only a few bits of data was extracted and no data on context or mechanisms | High rigour explains the methods and recruitment of care homes although does not explain the recruitment of residents. Also used a purposive approach to ensure that they got a range of different care home types but still may not be generalisable to all care homes due to small smaple size They used a evaluated audit tool and had a range of perspectives. |
| Jay 2014 | Moderately relevant looks like this could be the same study as Ilam 2018 a lot of the evidence is worded the same. Focus on one IPT but reports each resident separately, there is some outcomes context and mechanisms extracted | Moderate richness discusses one IPT in detail, can extract mechanisms, outcomes and context. | Moderate rigour. small sample size from only two care homes. No information how the care homes were recruited. Participants were randomly assigned to intervention but lead to only two participants being assigned to the landmark intervention. participants did not use both intervention types so individual differences could explain difference |
| Joy 2013 | Low Does mention wandering techniques but not in detail. No data extracted on context, mechanisms or outcomes. The information extracted same as other articles | Low- No data extracted on context mechanisms or outcomes. This is a commentary piece | This is a commentary piece so only views of the research and only mention techniques rather than discuss who they work for and why |
| Kearns 2007 | Moderate relevance this explores more about people opinions on strategies but some information about several IPTs | Moderate some data extracted for several IPTS but more about opinions on the strategies. There was some information on who the strategies work or do not work for | Moderate- small sample size. some key stakeholders were not interview so perspective may be skewed. Only one hospital so may not be generalisable |
| Lancioni 2011 | High. Quite a bit on intervention and mechanisms in relation to outcomes. Very little on context. although it is in a day centre, this information could be translated to care homes. | Moderate. Quantitative data relating to the intervention, only a few bits in the Discussion section relating to mechanisms. It discusses three participants in detail, enabling extraction of who the intervention may work for, although two of the participants had mild dementia. | Low. One institution, little details reported (doesn't look like it is residential?), only three participants, two of whom had mild dementia. Unclear how participants were recruited. Not clear how trustworthy the findings are. |
| Lancioni 2013 | Moderately relevant, builds on their work from 2011. Look at orientation but is in a day centre | Moderate richness - some detail relating to IPTs, discusses outcomes, context and some mechanism. The results and discussion is very short so richness is limited | Low. One institution, little details reported , only five participants, Unclear how participants were recruited. Not clear how trustworthy the findings are." |
| Lorey 2019 | High - related to a counter-argument about deceiving residents, with contexts and mechanisms | High - many examples with detail | Low - contextual factors unclear, methods for gathering arguments not reported on |
| Ludden 2019 | Moderate- Introduces a intervention for wayfinding that we have not extracted before but the information is limited | Moderate- information limited some outcomes, not context or mechanism | Low- case study, no information on the methodology for the cases studies, who did they observe, for how long. |
| MacAndrew 2019 | Moderate relevance, does talk about several IPTS but most of the paper was about preventing rather that supporting and did not fit in any IPTs and as it was a systematic review data that could be extracted was limited | Moderate as it was a systematic review data available to extract was limited some information on outcome but no infomation on context or mechanisms | Low- the systematic review was rigours with detailed systematic methods but the researchers reported that the papers included had very low quality methodology. Many of the papers were case studies. For many wandering was not the primary outcome and very few used validated measures |
| Marquardt 2009 | High. Relevant and possible to extract at least dyads. Elements of context and mechanism are presented in relation to outcome | High. Quantitative data, but there is an emphasis on explanation and quite a bit of detail, on one IPT. | Moderate. Little detail reported on the homes chosen, with little detail on recruitment and data collection and analysis. Difficult to tell if extraneous factors could have influenced raters' interpretation of orientation. Standardised scales were used, although there is no mention of where these came from. |
| Marquardt 2011 | Moderate- while the paper does review other studies so more data could be extracted from the individual papers this is relevant as it provides explanation and mechanisms for the results | Moderate- this is a review so limited information extracted about one specific study but does provide explanation for differences in results. Context, outcomes and mechanisms extracted | Moderate- this is a review so the rigour is dependent of the papers included. The paper does report setting and participants but no data on methods so unable to determine the rigour. |
| McGilton 2003 | High relevance- information extracted for IPT. Context, outcomes and some mechanisms extracted | Moderate- some information extracted that builds on other work in the review. Some context, outcomes and mechanisms extracted. | Moderate- RCT with one RA collecting outcome measures being blinded to the allocation. Only one care home was recruited. All residents had the opportunity to take part. No information on how the care home was recruited. |
| McQuilkin 2016 | Highl relevance, touches on several IPT with some outcomes and context extracted. | Moderate richness, discusses several IPTS with data extracted on outcomes and context. Little to no data on mechanisms. | Moderate rigour. Only looks at one care home with no discussion on recruitment of the care home. Small sample size with use of non-validated measures. used multiple methods to triangulate data. |
| Meiner 2000 | Low- only a few bits of data extracted more about preventing that supporting | Low- limited data extracted, no context, mechanisms or outcomes | Low- little information |
| Miskelly 2004 | Moderate - some detail on alerting staff (using the movement detection technology), however no information on what action staff then take, or even whether they would support or restrict wandering. | Moderate - some detail on how the system worked in practice and consideration of an ethical issue although not much detail overall. | Very little detail on methods, difficult to judge the rigour. Patient and staff perspectives not documented, pilot study with single group design. |
| Moore 2009 | Moderate relevance - some information on contexts and mechanisms, but very little detail on these and interventions | Low richness | Little detail on methods, not linked framework components to included studies |
| Neubauer 2018 | Moderate- discusses several IPTs | Moderate- Some context and outcomes, no mechanisms | Moderate- is a systematic reviews so determined on the rigour of the papers, high rigor for the systematic review process, assessed papers for methodology quality |
| Neubauer 2020 | Moderate- discusses people in the community so a lot was not relevant to care homes but some data extracted for IPTs and a lot of contextual information that other papers have not highlighted | Moderate- a lot of contextual information that other papers have not highlighted. limited to no outcomes or mechanism data | Moderate- A maximum variation sampling method was used, snowball sampling method was also used. The eligibility criteria such as English speaking only eliminated a lot of potential participants. wide range of view points including people with dementia |
| Neimeijer 2015 | Moderately relevant does talk about interventions to support wandering but more about opinions on using the technology rather than being able to extract COMC. One care home is for people with Intellectual disabilities who are younger so this may not be relevant | Moderate richness Some data extracted for several IPTs but little on context, outcome and mechanisms | Moderate rigour only two care homes but detailed recruitment process which seemed rigours. The researcher highlight that there may be questions about the reliability of the data. |
| O’Malley 2017 | Moderate- some information extracted on wayfinding but information limited some data extracted for mechanisms | Moderate- limited data, discussing other work in limited details, some information extracted on mechanisms | Low- no information on methodology of the studies and guidelines included, no information why or how the documents were found and selected |
| Olson 2021 | Moderate relevance the main topic of the paper is not wayfinding but wayfinding and access to garden spaces are explored | Moderate- as not the main topic there is only one section on wayfinding and some on access to garden space, but date extracted on outcomes and mechanisms and context | Low- Literature search so rigour is based on the documents included. Little information on the methods of the literature search or the papers included |
| Provencher 2008 | Moderately relevant, Discusses a technique for residents to learn routes | Moderate- Case study so goes into detail for context and outcomes, some on mechanisms from the theory | Low- Case study, no information on how the care home or participant was selected, the routes were short for fear of her getting lost so difficult to determine how the technique could be used more generally for wayfinding in care homes. |
| Rule 1992 | Moderate- some information on wayfinding with data extracted on context and mechanisms but information limited | Moderate- some information on wayfinding with data extracted on context and mechanisms but information limited. wandering was not the main topic of the paper | Low- this is from 1992 so must be considered with caution has the field has developed. There is no information on the rigour of the studies discussed and no information on how the studies were found forthe review. |
| Seetharaman 2022 | Moderate- more about person centred care but some relate to wandering | moderate- some information extracted on context, some on outcomes, none on mechanisms | Moderate- the data was from best practice guidelines in different countries, the rigour will be dependent on the guideline’s rigour |
| Shabha 2022 | High relevance- discusses wayfinding. Context and some mechanisms can be extracted | Moderate- a review so a lot of information but in-depth information about one thing is limited. A lot of context data extracted | Moderate- detailed description of how the review was conducted. They assessed the methodology quality of the papers but do not discuss the results of this in detail. They do comment on issues with rigour based the methodology of the papers included. Information on country of papers but no information on the settings or participants |
| Tufford 2018 | High relevance - possible to extract dyads and triads relating to the IPTs | Moderate richness - some detail relating to IPTs | Difficult to judge the rigour as not a great deal of detail on methods. Broad range of care homes and informants studied, range of perspectives |
| Van Buuren 2022 | Moderate relevance- some information extracted on wayfinding | Moderate- a lot of context information some mechanism, no outcomes | Moderate- detailed methodology, explanation on recruitment |
| Wiener 2021 | High relevance- discusses IPT6. Most of the information can be extracted from the papers they discuss but do bring some new information | High | Low. This is a commentary piece. |
| Wigg 2010 | Very relevant, compares a care home who support wandering and one who prevents wandering providing evidence for several IPTs for both supporting and counter arguments. Was able to extract context, and outcomes, there is some mention of mechanisms | High richness, a lot of data was extracted. There was data for outcomes, context and some information on mechanisms. covers several IPTs | Low to moderate. Difference in observation times one was ten years one was 6 months. they worked at one care home that could cause bias. No information about how the two care homes were recruited other than they worked at one. no great detail on the observation methods |
| Wigg 2020 | Moderate this has a lot of information for IPT 7 and 3 but it is mostly about GPS which we are not including. It is also more directed towards community living | Low- extracted some information but most not relevant and have not extracted any mechanisms, context. Some data extracted on outcomes and whom it works for | Little information on the methods to determine rigour. The rigour of the review would be determined by the rigour of the studies which are not discussed in this paper. |
| **Evidence sources identified through IPT-focused searches** | | | |
| Bautrant 2019 | Moderate- some information about wandering but paper more about BPSD as a whole | Low- some information but limited and difficult to decide if it fits with a IPT | Moderate- methodology limited- no information of recruitment of the care home |
| Bowes 2019 | High - information on contexts, mechanisms and outcomes in relation to the intervention is reported, relevant to a few PTs | Moderate - some detail on relevant interventions | High - rigorous process of identifying and selecting the data, inclusive criteria, broad search terms, large number of included studies, although the review does not focus directly on wandering |
| Dreyfus 2018 | High relevance discusses IPT 7 in great detail looking at the attitude of staff and family members. It is providing a new perspective where the individuals think that what they are doing is in the persons best intreast | Moderate- a lot of data extracted for outcomes, but little on mechanisms and context. | Moderate- Did look at 12 care homes but all from the same care organisation therefore care culture and policies could be the same which would impact attitude. The focus groups could have lead to people not being comfortable to voice their opinion although the participant types were seperated so that managers were not in with staff or family members. |
| Fleming 2010 | Moderate - a range of relevant interventions and some dyads might be extracted | Low - very little detail reported on the relevant findings | High - rigorous process of identifying and selecting the data, inclusive criteria |
| Li 2023 | Moderate relevance - IPT6 and some to IPT8 | Low richness - minimal data extracted | Low - unclear reporting of methods |
| Margot-Cattin 2006 | High - information on contexts, mechanisms and outcomes in relation to the intervention is reported, relavant to several IPTs | High - Rich data on how the intervention works and in which contexts | Ethnographic approach adds rigour thought triangulation of methods and staff and resident perspectives, however relative perspectives were not gathered and only one home was used |
| Mazzei 2014 | Moderate- some information relevant to IPT7 but limited information | Moderate- some context and outcomes, no mechanisms | Moderate- low sample size but expected due to setting, only one unit due to nature of study |
| Mikhaylova-O’Connell 2025 | High relevance- information extracted for several IPTs including some that are under-researched. | Moderate richness- data extracted for a number of IPTs, but not a huge amount of data extracted focuses on context, outcomes and mechanisms | Moderate- only looked at care homes in the north of the UK. the majority of participants were in senior leadership roles. Strong methodology with systematic analysis and reflexivity |
| Mueller 2013 | Moderate to high - information relating to the IPTs and possible to extract dyads however general view appears to be that wandering is problematic. | Moderate - some detail relating to some of the interventions and strategies, with alternate views at times | Moderate to high - Range of views of care workers and range of settings (and types of settings) sampled, although only three relatives. Observations taken as well as interviews. |
| Murphy 2017 | Low - minimal data related to wandering and nutrition/hydration, but there is little in this IPT | Low - some information extracted related | High - Clear reporting and transparency in qualitative methods |
| Murphy 2019 | Moderate- discusses IPT 10 which we have limited information on but only discuss strategies no context, outcome or mechanisms | Moderate- discusses IPT 10 which we have limited information on but only discuss strategies no context, outcome or mechanisms | Moderate- information from a organisation to create guidelines but no information on where the data came from |
| Neville 2006 | Moderate- discuss IPT 4 but information is limited | Low- limited data extracted no outcomes or mechanisms | Low. This is a commentary piece. |
| Padilla 2011 | Moderate - some relevant information, might be possible to extract dyads | Low - very little detail, summary of two studies | High - rigorous process of identifying and selecting the data, inclusive criteria, broad search terms, although the review does not focus directly on wandering |
| Padilla 2013 | Relevant - relates to the IPT and some dyads present. Should note that at times the marking intervention was paired with a cognitive behavioural intervention designed to reduce wandering by positively reinforcing other actions (and ignoring the resident when he made an exit attempt), which is not consistent with contemporary practice in the UK, nor with a pro-wandering stance. | Moderate - some detail on speculated mechanisms although findings not very detailed | Low - only looked at one patient. Method was scientific, but nevertheless the findings may have been different in other participants in other settings - it is difficult to know from this study. |
| Passini 2000 | High- this discusses a number of IPTs, it adds new information and is the paper that many of the other included studies discuss | High- detailed data extraction including context, outcomes and mechanisms | Moderate- detailed methodology, some explanation for recruitment although may not be representative as only one care home and small sample size |
| Roberts 1999 | Moderate - strategies to address entering unsafe spaces, with some context/mechanism information although not much | Low - very little detail reported | Low - study type unclear, setting largely unclear, very little detail on methods. Looks like one setting. |
| Tseng 2022 | Moderate - relevant to IPT6 | Low - little data extracted | Low - small sample size, one prototype |
| Van Hoof 2013 | Moderate- community based, only limited information on wandering | Low- only a few bits extracted on wandering, little context, no outcomes or mechanisms | Moderate- limited information on methodology |
| Van Liempd 2023 | Moderate relevance - little on mechanism, descriptive data about outcomes | Low richness - data extracted with a lot of overlap | High rigour - transparent methods, use of framework to guide analysis and reporting |
| **Grey literature searching** | | | |
| Anonymous 2018 | Low- only one piece of data extracted little detail | Low- only one line extracted, no data on context, outcomes or mechanisms | Low- no information |
| Britton 2021 | Moderate- some strategies but data extracted limited but on a IPT that we have very limited data on | Moderate- data extracted for several IPT but only discuss strategies no context, outcomes or mechanisms | Low- commentary piece difficult to assess the rigour of it |
| Sharp 2021 | Moderate- only one piece of data extracted but on a IPT that we have very limited data on | Moderate- only one piece of data extracted but on a IPT that we have very limited data on | Low- commentary piece but is from a registered nurse and organisation that deals with nutrition although difficult to assess the rigour of it |
