## Supplement 3 for "Strategies to support safe wandering in care homes for older adults – what works, for whom, and in which circumstances?: A realist synthesis"

Supplement 3: Evidence mapping

### Table S3.1: Programme theory evidence mapping

|  | **Initial programme theories** | | | | | | | **Refined programme theories** | | | | |
| --- | --- | --- | --- | --- | --- | --- | --- | --- | --- | --- | --- | --- |
| **First author, date** | **IPT3** | **IPT4** | **IPT6** | **IPT7** | **IPT8** | **IPT9** | **IPT10** | **Personalised care** | **Monitoring** | **Navigation** | **Managing access** | **Hydration & nutrition** |
| **Evidence sources identified through initial database searches** | | | | | | | | | | | | |
| Alam 2023 | ✔ |  | ✔ | ✔ |  |  |  |  | ✔ | ✔ |  |  |
| Anderson 2023 |  |  | ✔ | ✔ | ✔ |  |  |  |  |  | ✔ |  |
| Anonymous 2007 | ✔ |  | ✔ | ✔ |  |  |  |  | ✔ | ✔ | ✔ |  |
| Apple 2015 | ✔ |  | ✔ | ✔ | ✔ | ✔ |  |  | ✔ |  | ✔ |  |
| Barrett 2020 | ✔ | ✔ | ✔ | ✔ | ✔ | ✔ |  | ✔ | ✔ |  | ✔ |  |
| Benbow 2017 | ✔ |  | ✔ | ✔ | ✔ |  |  | ✔ | ✔ |  | ✔ |  |
| Brush 2008 |  |  | ✔ |  |  |  |  |  |  | ✔ |  |  |
| Brush 2015 |  |  | ✔ |  |  |  |  |  |  | ✔ |  |  |
| Caffo 2014 |  |  | ✔ |  |  |  |  |  |  | ✔ |  |  |
| Cheung 2021 | ✔ |  |  | ✔ |  |  |  |  | ✔ |  |  |  |
| Cohen-Mansfield 1998 |  |  |  |  | ✔ |  |  |  |  |  | ✔ |  |
| Detweiler 2008 |  |  |  | ✔ | ✔ | ✔ |  |  |  |  |  |  |
| Detweiler 2009 |  |  |  |  | ✔ |  |  |  |  |  | ✔ |  |
| Dewing 2011 |  | ✔ |  | ✔ |  | ✔ |  | ✔ |  | ✔ |  |  |
| Dickinson 1998 | ✔ | ✔ |  | ✔ |  |  |  | ✔ | ✔ |  | ✔ |  |
| Dickinson 1995 |  |  |  | ✔ |  |  |  |  |  |  | ✔ |  |
| Duffy 2019 | ✔ | ✔ |  | ✔ | ✔ |  |  | ✔ | ✔ |  | ✔ |  |
| Faith 2014 | ✔ | ✔ | ✔ | ✔ | ✔ | ✔ |  | ✔ | ✔ | ✔ | ✔ |  |
| Fine 2015 |  |  | ✔ |  | ✔ |  |  |  |  |  | ✔ |  |
| Gibson 2004 |  |  | ✔ |  |  |  |  |  |  |  | ✔ |  |
| Griffiths 2024 | ✔ | ✔ | ✔ | ✔ | ✔ | ✔ |  | ✔ |  | ✔ | ✔ |  |
| Gulwadi 2013 |  |  | ✔ |  |  |  |  |  |  |  |  |  |
| Heward 2022 |  | ✔ | ✔ |  | ✔ | ✔ |  | ✔ |  | ✔ | ✔ |  |
| Hirst 1989 |  | ✔ |  |  |  |  | ✔ | ✔ |  |  |  | ✔ |
| Holthe 2018 | ✔ |  | ✔ |  |  |  |  |  | ✔ |  |  |  |
| Hussain 1987 |  |  |  | ✔ |  |  |  |  |  |  | ✔ |  |
| Ilem 2018 |  |  | ✔ |  |  |  |  |  |  | ✔ |  |  |
| Innes 2011 |  |  | ✔ | ✔ | ✔ |  |  |  |  | ✔ | ✔ |  |
| Jay 2014 |  |  | ✔ |  |  |  |  |  |  | ✔ |  |  |
| Joy 2013 |  |  | ✔ | ✔ |  |  |  |  |  | ✔ | ✔ |  |
| Kearns 2007 | ✔ |  |  | ✔ |  |  |  |  | ✔ |  | ✔ |  |
| Lancioni 2011 |  |  | ✔ |  |  |  |  |  |  | ✔ |  |  |
| Lancioni 2013 |  |  | ✔ |  |  |  |  |  |  |  |  |  |
| Lorey 2019 |  |  |  | ✔ |  |  |  |  |  |  | ✔ |  |
| Ludden 2019 |  |  | ✔ |  |  |  |  |  |  | ✔ |  |  |
| MacAndrew 2019 | ✔ |  | ✔ | ✔ |  |  |  |  | ✔ |  |  |  |
| Marquardt 2009 |  |  | ✔ |  |  |  |  |  |  | ✔ |  |  |
| Marquardt 2011 |  |  | ✔ | ✔ |  |  |  |  |  | ✔ | ✔ |  |
| McGilton 2003 |  |  | ✔ |  |  |  |  |  |  | ✔ |  |  |
| McQuilkin 2016 | ✔ | ✔ | ✔ | ✔ | ✔ | ✔ |  | ✔ | ✔ | ✔ | ✔ |  |
| Meiner 2000 | ✔ | ✔ |  | ✔ |  | ✔ |  |  |  | ✔ |  |  |
| Miskelly 2004 | ✔ |  |  | ✔ |  |  |  |  | ✔ |  |  |  |
| Moore 2009 | ✔ |  | ✔ | ✔ | ✔ |  |  |  | ✔ |  | ✔ |  |
| Neubauer 2018 | ✔ |  | ✔ | ✔ | ✔ |  |  |  | ✔ |  | ✔ |  |
| Neubauer 2020 | ✔ |  | ✔ | ✔ |  |  |  |  | ✔ |  |  |  |
| Neimeijer 2015 | ✔ |  |  | ✔ | ✔ | ✔ |  |  | ✔ | ✔ | ✔ |  |
| O’Malley 2017 |  |  | ✔ |  |  |  |  |  |  | ✔ |  |  |
| Olson 2021 |  |  | ✔ |  | ✔ |  |  |  |  | ✔ | ✔ |  |
| Provencher 2008 |  |  | ✔ |  |  |  |  |  |  | ✔ |  |  |
| Rule 1992 |  |  | ✔ |  |  |  |  |  |  | ✔ |  |  |
| Seetharaman 2022 | ✔ | ✔ | ✔ | ✔ | ✔ |  |  | ✔ | ✔ | ✔ | ✔ |  |
| Shabha 2022 | ✔ |  | ✔ |  |  |  |  |  |  | ✔ | ✔ |  |
| Tufford 2018 |  |  |  | ✔ | ✔ | ✔ |  |  |  |  | ✔ |  |
| Van Buuren 2022 |  |  | ✔ |  |  |  |  |  |  | ✔ |  |  |
| Wiener 2021 |  |  | ✔ |  |  |  |  |  |  | ✔ |  |  |
| Wigg 2010 | ✔ | ✔ |  | ✔ | ✔ | ✔ |  | ✔ | ✔ |  | ✔ |  |
| Wigg 2020 | ✔ |  |  | ✔ | ✔ |  |  |  | ✔ |  |  |  |
| **Evidence sources identified through IPT-focused searches** | | | | | | | | | | | | |
| Bautrant 2019 |  |  | ✔ |  |  |  |  |  |  | ✔ |  |  |
| Bowes 2019 |  |  | ✔ | ✔ | ✔ |  |  |  | ✔ |  | ✔ |  |
| Dreyfus 2018 |  |  | ✔ | ✔ | ✔ |  |  |  |  |  | ✔ |  |
| Fleming 2010 |  |  | ✔ | ✔ |  |  |  |  |  |  | ✔ |  |
| Li 2023 |  |  | ✔ |  | ✔ |  |  |  |  |  | ✔ |  |
| Margot-Cattin 2006 | ✔ |  | ✔ | ✔ | ✔ |  |  |  | ✔ | ✔ | ✔ |  |
| Mazzei 2014 |  |  |  | ✔ | ✔ |  |  |  |  | ✔ | ✔ |  |
| Mikhaylova-O’Connell 2025 | ✔ | ✔ |  |  |  | ✔ | ✔ | ✔ | ✔ |  |  | ✔ |
| Mueller 2013 | ✔ |  | ✔ | ✔ |  |  |  |  |  |  | ✔ |  |
| Murphy 2017 |  |  |  |  |  |  | ✔ |  |  |  |  | ✔ |
| Murphy 2019 |  |  |  |  |  |  | ✔ |  |  |  |  | ✔ |
| Neville 2006 | ✔ | ✔ |  |  |  |  |  | ✔ |  |  |  |  |
| Padilla 2011 |  |  |  | ✔ |  |  |  |  |  | ✔ | ✔ |  |
| Padilla 2013 |  |  |  | ✔ |  |  |  |  |  |  | ✔ |  |
| Passini 2000 |  |  | ✔ | ✔ | ✔ | ✔ |  |  | ✔ | ✔ | ✔ |  |
| Roberts 1999 |  |  |  | ✔ |  |  |  |  |  |  | ✔ |  |
| Tseng 2022 | ✔ |  | ✔ |  |  |  |  |  |  | ✔ |  |  |
| Van Hoof 2013 | ✔ |  | ✔ | ✔ | ✔ |  |  |  |  | ✔ |  |  |
| Van Liempd 2023 |  |  |  |  | ✔ |  |  |  |  | ✔ | ✔ |  |
| **Grey literature searching** | | | | | | | | | | | | |
| Anonymous 2018 |  |  | ✔ |  |  |  |  |  |  | ✔ |  |  |
| Britton 2021 |  |  |  |  | ✔ | ✔ | ✔ |  |  |  | ✔ |  |
| Sharp 2021 |  |  |  |  |  |  | ✔ |  |  |  |  | ✔ |

*Black ticks denote contribution; grey ticks denote minor contribution*
